# Machine learning-based analysis of genomic and transcriptomic data unveils sarcoma clusters with superlative prognostic and predictive value

**DOI:** 10.1101/2025.01.31.25321492

**Authors:** Miguel Esperança-Martins, Hugo Vasques, Manuel Sokolov Ravasqueira, Maria Manuel Lemos, Filipa Fonseca, Diogo Coutinho, Jorge Antonio López, Richard S.P. Huang, Sérgio Dias, Lina Gallego-Paez, Luís Costa, Nuno Abecasis, Emanuel Gonçalves, Isabel Fernandes

## Abstract

Soft tissue sarcomas (STS) histopathological classification system has several conceptual caveats, impacting prognostication and treatment. The clinical and molecular-based tools currently employed to estimate prognosis also have limitations. Clinically driven molecular profiling studies may cover these gaps. We performed DNA sequencing (DNAseq) and RNA sequencing (RNAseq), portraying the molecular profile of 102 samples of 3 of the most common STS subtypes. The RNAseq data was analyzed using unsupervised machine learning models, unravelling previously unknown molecular patterns and identifying 4 well-defined transcriptomic clusters. These transcriptomic clusters have a clear prognostic value, a finding that was externally validated. This transcriptomic cluster-based classification’s prognostic value is superior to the prognostic accuracy of currently used clinical-based (SARCULATOR nomograms) and molecular-based (CINSARC) prognostication tools. The analysis of DNAseq data from the same cohort of samples revealed a plethora of unique and, in some cases, never documented molecular targets for precision treatment across different transcriptomic clusters.

## Introduction

Sarcomas are, biologically, rooted deeply in vertebrates evolutionary history, as consubstantiated in a paleopathology study that documented findings compatible with an osteosarcoma on the femur of a 240-million-year-old stem-turtle from the Triassic period [1]. Even though sarcomas are not modern vertebrate and human physiological defects or recently discovered pathological entities, the poor descriptive depth and the immutability of the attributes that are used to portray them throughout the last decades reflect our scarce comprehension of its biology.

Sarcomas are almost invariably characterized as a group of rare and heterogenous mesenchymal malignancies [2,3,4]. Sarcomas’ heterogeneity is, conceptually, mainly a product of the currently applied histopathological classification system. This system fragments sarcomas in 50 to 150 histological subtypes, with approximately 20% of them being defined as “ultra-rare”, with an incidence of less than 1 in 1.000.000 [5].

Sarcomas histopathological classification system has important limitations. Firstly, this system is eminently morphological and based on the resemblance of neoplastic tissue to the type of normal tissue counterpart (i.e., line of differentiation) [6,7]. Methodologically, this approach hinged on seeking morphological similarities between the neoplastic tissue and different types of normal tissues is indirect and non-specific. Moreover, this system is intrinsically complex and prone to errors, as it is illustrated by reported sarcomas’ overall diagnostic discrepancy rates of 28.2-56% and major diagnostic discrepancy rates (mainly due to discordances in histological types and grades) of 16.4-37% between referring and tertiary reference centers in different series [8–13].

This imperfect histopathological classification system shapes soft tissue sarcomas (STS) management by impacting both retroperitoneal (RPS) and extremity (eSTS) STS prognosis estimation and respective prognostication accuracy. The clinical nomograms that are most widely used for prognostication purposes, available at the SARCULATOR application, incorporate STS histopathological subtype along with other histopathological variables as critical and defining factors. Besides this, although sarcoma’s treatment approach is largely based on a “fit-for-all” principle, there are specific treatment strategies for certain subtypes, which means that the above-mentioned diagnostic inaccuracies may directly impact treatment decisions. In some series, the modification of the histopathological diagnosis led to the modification of treatment strategy (surgical treatment, medical treatment and both) in up to 15% of cases [13]. Furthermore, adding outstanding fragmentation and heterogeneity to sarcomas’ rarity creates a particularly deleterious context for pre-clinical studies and for early and late-phase clinical trials development (specially in what recruitment and design is concerned), hampering drug discovery and development in sarcomas. The negative influence of the currently employed classification method on drug discovery and development also derives from this system’s static cytoarchitectural criteria, which do not fully capture the fluidity and dynamicity of the profusion of unique molecular landscapes of different sarcomas that are now being brought to light.

Molecular-based approaches and tools may fill in some of the conceptual gaps of the histopathological classification system. In fact, the use of molecular analytical instruments, such as comprehensive genomic profiling, modifies clinical diagnoses in a significant percentage of neoplasms suspicious of sarcoma and impacts their management, as reported in a recent study [14]. In this study, whole-transcriptome sequencing led to the reclassification of 7% of the histopathological diagnoses and found treatment relevant variants in 15% of cases [14]. Other studies have reported even higher diagnostic revision rates - between 3 and 14% - with the use of prospective genome-wide profiling [15,16,17]. Some of these broad sequencing studies also revealed the presence of actionable molecular alterations in higher percentages of STS patients – 31.7% -, while some real-world series identified druggable molecular alterations in 37.2% of STS patients, leading to the prescription of personalized treatment according to the identified molecular alterations in 31.2% of patients [18].

In the same way, a plethora of distinct molecular-based approaches (both single and multi-omics) are currently showing previously unknown angles of sarcomagenesis, permitting a more granular cartography of the pivotal molecular alterations that characterize different sarcomas, and are subsequently allowing an increment of prognosis estimation accuracy. Molecular prognostic and predictive biomarkers, such as genomic and transcriptomic signatures (of which the Complexity INdex in SARComas (CINSARC) and Genomic Grade Index (CGI) are good examples), together with proteomic and metabolomic fingerprints, are starting to pave the way for accurate prognosis definition and personalized treatment approaches identification in sarcomas [19–23].

The classification system should evolve from a crystalized and architectural archetype to a dynamic mesh that is capable to capture and comprise both common molecular drivers and specific molecular adaptations, allowing researchers to better estimate prognosis (overcoming clinical-based nomograms that integrate histopathological features such as SARCULATOR [24], gene expression-based signatures eminently related to mitosis and chromosome integrity, such as CINSARC [25], and even the combination of these prognostication instruments, embodied by CINSARCULATOR [26]), to more accurately find biological patterns and to better design studies and trials focused on tackling molecular alterations of specific sarcomas.

In order to achieve this, we identified, from the analysis of different STS samples, specific molecular signatures and transcriptomic clusters that correlate with clinical outcomes, showing impressive prognostic value, and that may guide selective personalized treatment strategies, displaying a potential predictive utility.

## Results

### Clinical characteristics of the study cohort

The 102 samples that were used for this study were obtained from 101 patients. These 101 patients displayed a median age of 67 (IQR 19.7) years-old, and a balanced gender distribution (50.5% male) (**Supplementary Table 1**). Twenty-five (25) patients had a diagnosis of dedifferentiated liposarcoma (DDLPS), 25 patients had a diagnosis of leiomyosarcoma (LMS), and 51 patients had a diagnosis of undifferentiated pleomorphic sarcoma (UPS). These patients STS’s were predominantly located in the lower limb (N=49, 48.5%), followed by the retroperitoneum (N=31, 30.5%). The primary malignant tumors (sarcomas) had a median size of 13 (IQR 10.0) cm. Two (6.5%) of the 31 retroperitoneal sarcomas were multifocal. All of these 101 patients samples were high-grade (Grade 3). The great majority of cases (N=96, 95%) presented with localized disease. Five (5%) patients were metastatic at diagnosis. Among these 5 patients, 4 (80%) had lung metastases and 1 (20%) had ganglionic mediastinal metastases. Three of these five patients (60%) were submitted to surgery with a palliative intent and 2 (40%) were not surgically interventioned.

Surgery was the most frequently employed treatment strategy (N=99, 98.0%). Among the 99 patients that were surgically interventioned, 2 (2%) had already been operated in another institution. From the 97 patients that were submitted to surgery by the IPOLFG surgical team, 94 (96.9%) were operated with a curative intent and 3 (3.1%) with a palliative intent. Among the 94 patients that were operated with curative intent, 2 (2.1%) had sarcomas that were deemed unresectable during surgery and 92 (97.9%) had resectable disease (**Supplementary Table 2**). The resection margin status was R0/R1 in 96.7% (N=89) of cases. Among the 94 patients that were operated with a curative intent, 3 (3.2%) were submitted to neoadjuvant treatment - 2 were treated with neoadjuvant chemotherapy (a doxorubicin-ifosfamide-based regimen was used in both cases) and 1 with neoadjuvant external radiotherapy (50 Gy/25 fractions). Fifty-eight (63.0%) patients received adjuvant treatment, primarily external radiotherapy (N=55, 94.8%).

Three patients with a R2 resection were excluded from the pool of locally recurrent cases since they were considered to have persistent disease.

Different oncological outcomes were evaluated for the patients who were submitted to a resection with curative intent (N=94), and whose resection margins were R0/R1 (N=89, 94.7%), during a median follow-up period of 27 (IQR 51.3) months since their diagnose. Of these 89 patients, 29 (32.6%) had already had a previous local treatment in other institution (surgery or radiotherapy). Among these 89 patients, the local recurrence rate was 46.1% (N=41), with a median time to local recurrence of 14 (IQR 29.0) months. Among the 96 patients without distant metastasis *ab initio*, the distant metastasis rate was 41.7 % (N=40). Metastases were mostly found in the lungs (N=34, 85%), with a median time to distant metastasis of 13 (IQR 17.2) months. The metastasis-free survival (MFS) rate during the follow-up period was 34.4% (N=33) and OS rate during the follow-up period for these patients was 43.8% (N= 42) with a median follow-up of 27 (IQR 51.5) months. The 5-year MFS rate and the 5-year OS rate for these patients were 37% and 46%, respectively. When all the patients are considered (N=101), the OS rate during the follow-up period was 42.6% (N=43), with a median follow-up of 25 (IQR 51.9) months. The 5-year OS rate for all the 101 patients was 44%.

### Analysis of RNAseq data using unsupervised machine learning methods, namely consensus clustering, identified 4 transcriptomic clusters with distinctive molecular signatures

Data from 74 samples (16 DDLPS, 15 LMS and 43 UPS) of 74 patients was considered for the RNAseq analysis (**see Methods**).

Transcriptomics consensus clustering identified 4 transcriptomic clusters (the optimal number of clusters was found using the Elbow method) **(Supplementary Figures 1a and 1b)**. Each transcriptomic cluster is portrayed by differential expression, either over or under expression, of a certain plethora of genes and of associated pathways (**Figure 1a, Figure 1b, Supplementary Figure 2** and **Table 1**).

**Figure 1.**
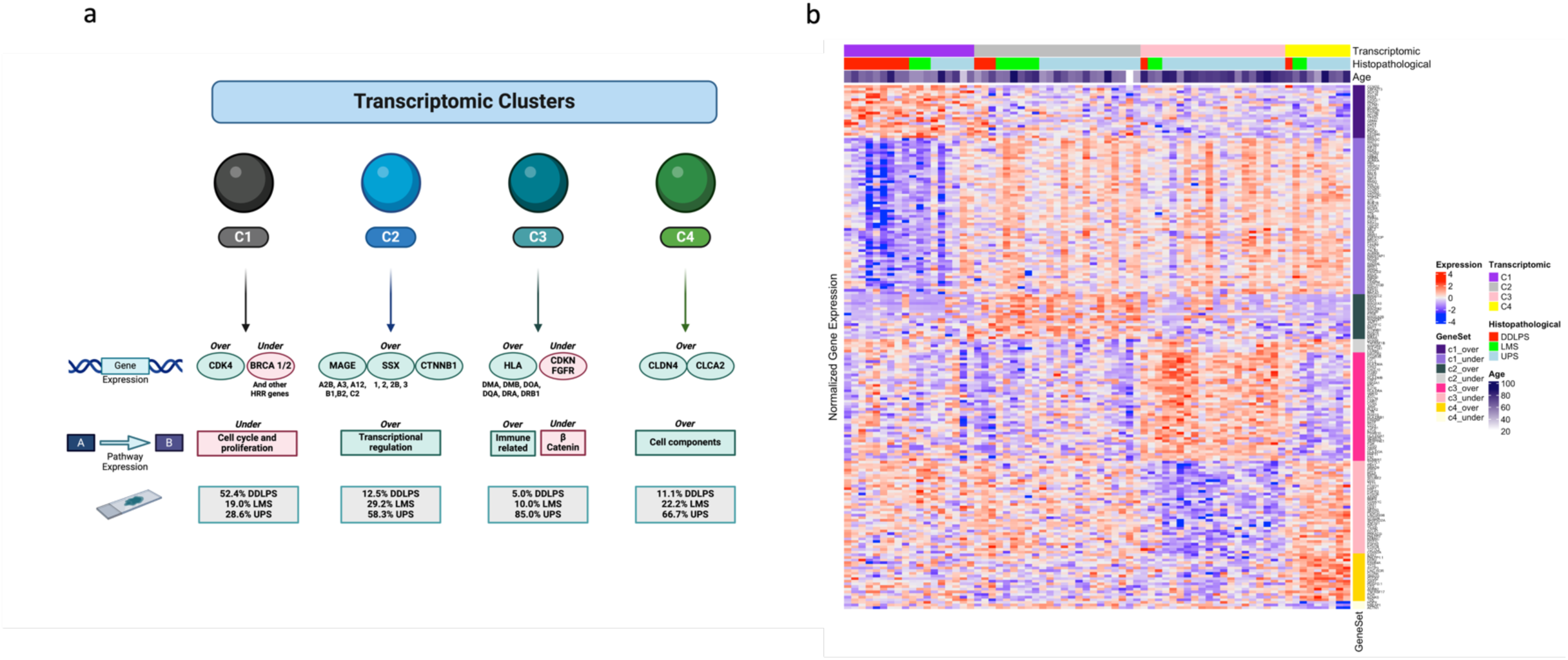
– The four identified transcriptomic clusters and their defining molecular features. **a)** Schematic of the genes and pathways whose expression pattern most distinctively portray each cluster and of the histopathological subtypes that differentially compose each cluster. Created in BioRender. Esperança-Martins, M. (2025) https://BioRender.com/c93w707 **b)** Heatmap plot displaying clinical and molecular (normalized gene expression data) features of each transcriptomic cluster.

**Table 1.**
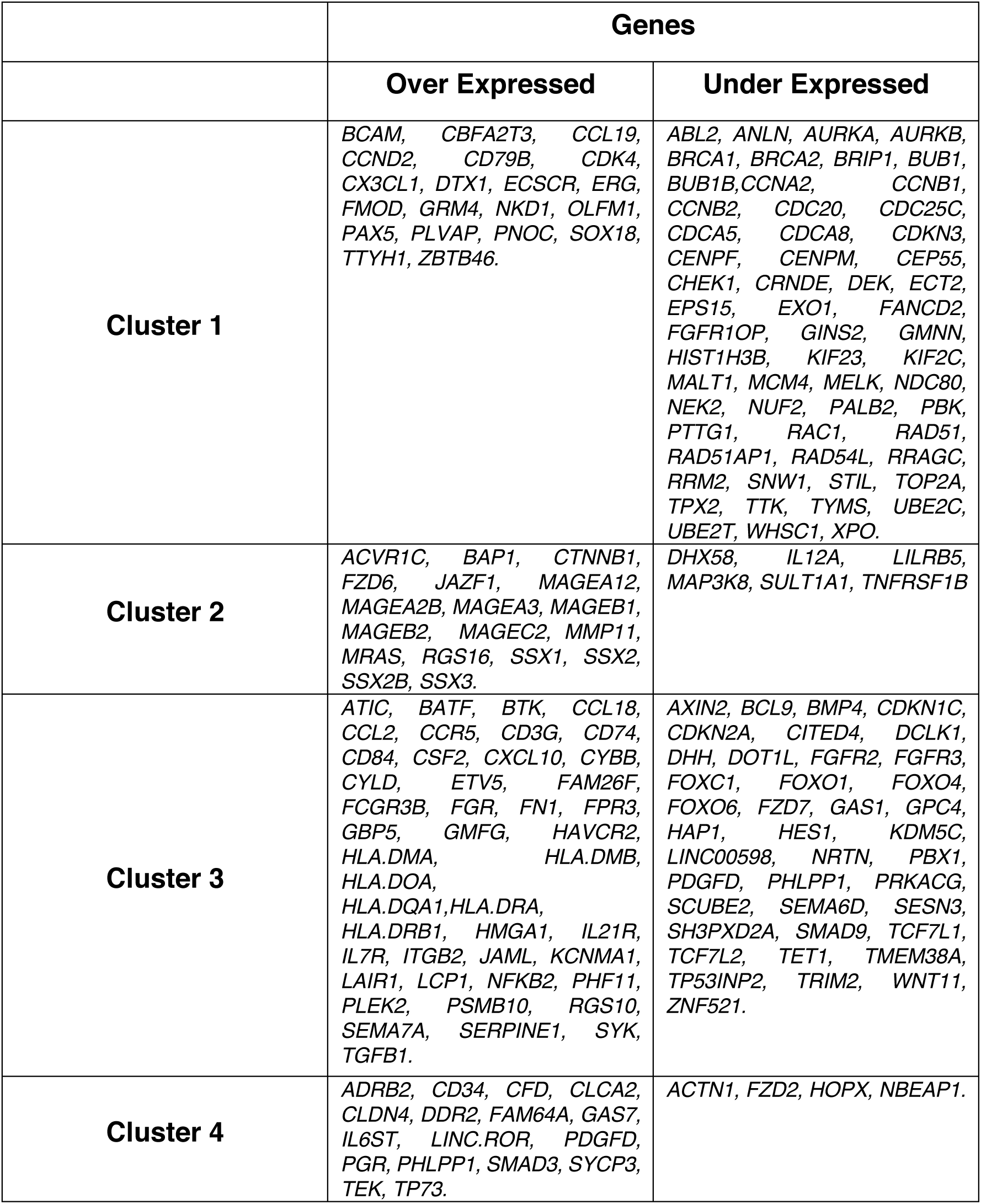
–Distribution of differentially expressed genes (over and under expressed) per transcriptomic cluster.

For each cluster, the differential gene expression analysis and subsequential KEGG pathway enrichment analysis were used to portray the cluster individual molecular landscape.

<u>Cluster 1 (C1)</u> is more distinctively portrayed by the over expression of genes that encode cyclin-dependent kinases and cyclins such as *CDK4* and *CCND2*. This cluster is also characterized by the over expression of genes that encode chemokines and transcription factors and by the under expression of an impressive array of genes that are involved in DNA homologous recombination repair (HRR) mechanisms, such as *BRCA1, BRCA2, FANCD2, PALB2, RAD51, CHEK1*, and *BRIP1*. Globally, there is an under expression of cell cycle and proliferation pathways (probably associated with the under expression of a significant number of genes involved in HRR and of a number of genes encoding cyclins, other than CCND2, as it is shown in **Table 1**). This cluster is mainly composed by samples that were classified, according to the currently used histopathological classification, as DDLPS (52.4%), with UPS (28.6%) and LMS (19.0%) samples also being integrated in this cluster.

<u>Cluster 2 (C2)</u> is predominantly defined by the over expression of different cancer testis antigens (CTA) genes, namely a plethora of MAGE genes (such as *MAGEA2B, MAGEA3, MAGEA12, MAGEB1, MAGEB2* and *MAGEC2*), and different SSX genes (such as *SSX1, SSX2, SSX2B* and *SSX3*). The over expression of *CTNNB1* is also verified in this cluster. It is important to emphasize that our study cohort included DDLPS, LMS and UPS samples, not comprising either synovial sarcoma or myxoid/round cell liposarcoma samples. There is an over expression of transcriptional regulation pathways in this cluster. This cluster is mostly composed by samples classified as UPS (58.3%), with LMS (29.2%) and DDLPS (12.5%) samples also being represented in this specific cluster.

Cluster 3 (C3) is specifically characterized by the over expression of genes that encode Major Histocompatibility Complex (MHC) class II/ Human Leukocyte Antigen (HLA) class II (*HLA-DMA, HLA-DMB, HLA-DOA, HLA-DQA, HLA-DRA* and *HLA-DRB1*) genes. Besides HLA class II genes, an over expression of *TGFβ1, ETV5, BTK* and *BATF* genes is also verified. On the other hand, the under expression of CDKN (*CDKN1C* and *CDKN2A*) and FGFR (*FGFR 2* and *FGFR3*) genes also characterize this cluster. In terms of pathways, this cluster is marked by an over expression of immune related pathways and an under expression of the β-catenin pathway. Samples labelled as UPS (85.0%) are predominant, while LMS (10.0%) and DDLPS (5.0%) samples are also integrated in this cluster.

<u>Cluster 4 (C4)</u> is represented by the over expression of a plethora of genes that encode different structural protein elements, such as claudin (*CLDN*) 4 (this gene encodes a membrane protein that is a component of epithelial cell tight junctions), *CLCA 2*, and *GAS 7*. Besides this, there is also an over expression of other genes such as *SMAD3* and *PDGFD*. Interestingly, an under expression of *ACTN1* is verified. There is an overall over expression of cell components pathways. This cluster is also principally composed by UPS (66.7%) samples, moreover, incorporating both LMS (22.2%) and DDLPS (11.1%) samples.

### The identified molecular signatures and respective transcriptomic clusters have a clear prognostic value, a finding that was externally validated

The newly identified transcriptomic clusters were included, alongside other key demographical, clinical, and histopathological data in the pool of variables that were considered for analysis using a Cox Proportional Hazards Model to estimate and compare the differential impact of each of these variables on OS (using the study cohort). This analysis revealed that clusters C2, C3, and C4 are negative prognostic factors. Specifically, the hazard ratios (HR) that were found were: C2 (HR 5.10; 95% CI 1.81-14.34; P=0.002), C3 (HR 4.47; 95% CI 1.39-14.45; P=0.01), and C4 (HR 7.66; 95% CI 2.06-28.53; P=0.002) (Figure 2a). An Analysis of Variance (ANOVA) test was applied to the Cox Proportional Hazards Model and demonstrated that transcriptomic clusters were the variable with the most significant correlation with OS (P<0.01) (Figure 2b).

**Figure 2.**
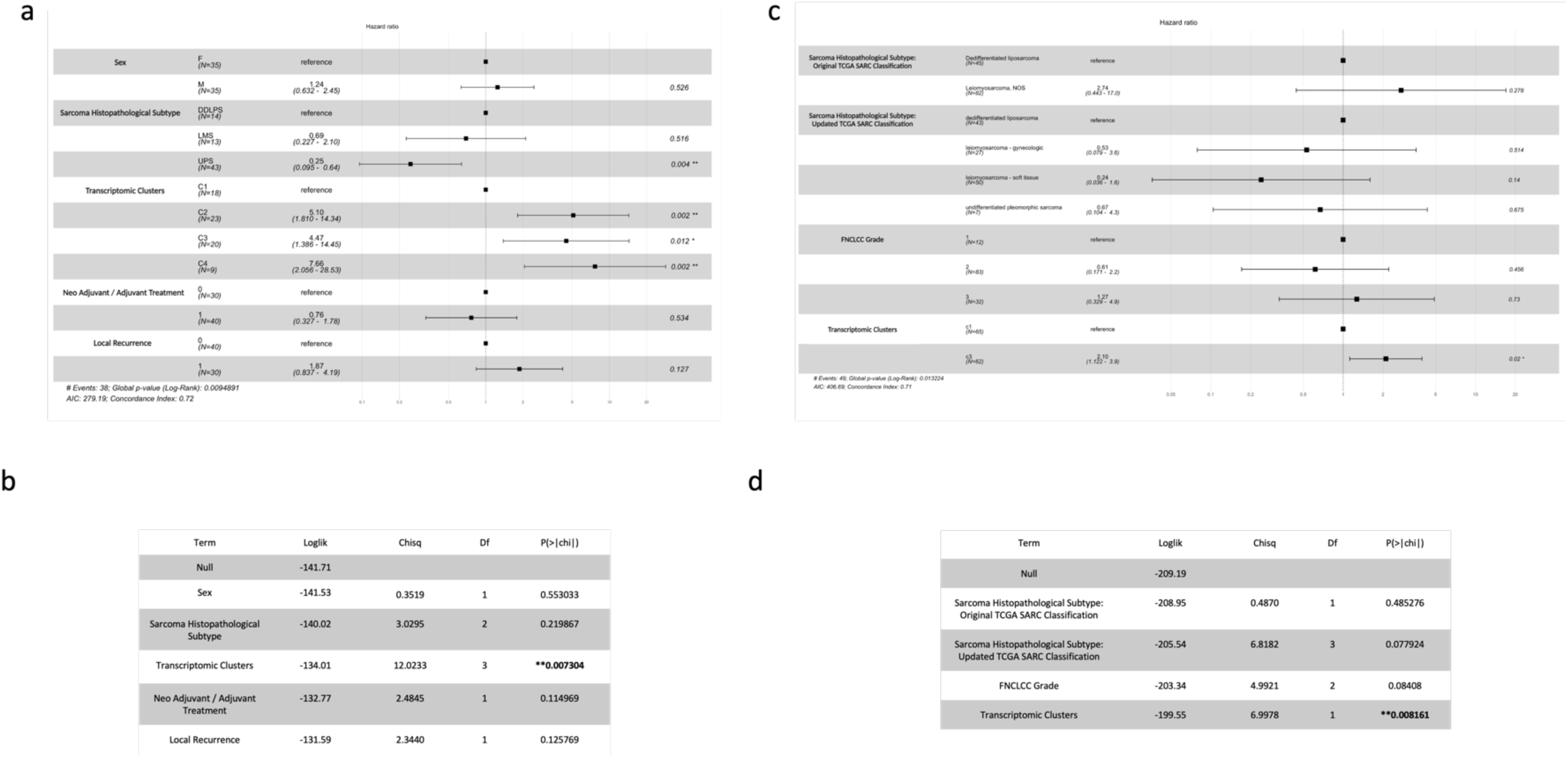
– Transcriptomic clusters and their respective molecular signatures exquisite prognostic value. **a)** Forest plot showing the results of the evaluation of the differential impact of distinct demographical, clinical, histopathological and molecular variables on OS in the study cohort using a Cox Proportional Hazards Model. **b)** Table displaying the results of the ANOVA test applied to the Cox Proportional Hazards Model to assess the predictive ability of different variables for OS estimation in the study cohort. **c)** Forest plot showing the results of the evaluation of the differential impact of distinct histopathological and molecular variables on OS considering the TCGA-SARC patients (classified in accordance with the transcriptomic clusters-based classification) using a Cox Proportional Hazards Model. **d)** Table displaying the results of the ANOVA test applied to the Cox Proportional Hazards Model to assess the predictive ability of different variables for OS estimation in the validation cohort (TCGA-SARC).

To externally validate these findings, we used the TCGA-SARC dataset, namely the data of patients with the same STS histopathological subtypes than the patients included in our study cohort (DDLPS, LMS and UPS) (N=127). We employed normalized gene expression data to reclassify patients into our transcriptomic cluster-specific gene signatures using single-sample Gene Set Enrichment Analysis (ssGSEA) (**see Methods**). Patients were assigned to the transcriptomic cluster with the lowest significant FDR-adjusted p-value. This led to the classification of these TCGA-SARC patients either into C1 (N=65) or C3 (N=62). Accordingly, a significant enrichment of the TCGA-SARC patientś samples to C1_under and C3_over was verified (**Supplementary Figure 3**). An analysis using a Cox Proportional Hazards Model was carried out, incorporating histopathological variables, such as the histopathological classification that was originally used in TCGA-SARC (that grouped LMS and UPS together), a recently proposed histopathological classification that distinguishes gynecological LMS, soft tissue LMS and UPS (and which is currently used for patient stratification), and the FNCLCC grade, and the transcriptomic clusters. This analysis confirmed that C3-enriched patients have a worse prognosis (HR 2.28; 95% CI 1.19-3.9; P=0.02) (Figure 2c). An ANOVA test of the Cox Proportional Hazards Model showed, once again, that the transcriptomic cluster-based classification was the most significant predictor of overall survival (P<0.01) (Figure 2d).

### The enrichment in C1 and C3 of the TCGA-SARC population and the statistical significance of the correlation between transcriptomic clusters and survival persist even when UPS patients are excluded

As described in Methods, using the TCGA-SARC dataset, we evaluated if the removal of UPS patients from the patients pool would alter the previously verified molecular enrichment of these STS patients samples in C1 and C3 (C1_under; C3_over) and modify the previously verified statistically significant correlation between the transcriptomic clusters-based classification and OS. When UPS patients were removed from the considered TCGA-SARC patients population, the molecular enrichment of the population in C1 and C3 (C1_under; C3_over) (**Supplementary Figure 4**) and the correlation between the transcriptomic clusters-based classification and OS remained statistically significant (namely the correlation between C3 and OS) (**Supplementary Figure 5**).

However, we would like to underline that within our study population, UPS was the most frequently represented STS histopathological subtype (N=51 (of 101), 50.4%), while on the TCGA-SARC population (considering the distribution per STS histopathological subtype displayed on the reviewed histopathological classification of TCGA-SARC), UPS was the less frequently represented STS histopathological subtype (N=7 (of 127), 5.5%).

### A molecular signature and transcriptomic cluster-based classification outperforms the SARCULATOR clinical nomograms in terms of prognostic value

We conducted a comparative analysis between the prognostic values of a molecular signature/transcriptomic cluster-based classification and the clinical nomograms available at SARCULATOR (SARCULATOR). 67 patients within the study cohort had their 5-year OS probability estimated following the use of SARCULATOR nomograms (**see Methods**). The median 5-year predicted OS was 57% (IQR 26.5%).

C-indexes of the different Cox Proportional Hazard models for OS were calculated and then compared (**see Methods**). The following models were considered: SARCULATOR 5-year OS prediction (SARC); transcriptomic clusters (TC); SARCULATOR 5-year OS prediction combined with transcriptomic clusters (SARC+TC); and finally, transcriptomic clusters combined with age (TC+AGE).

The TC+AGE model showed the strongest OS predictive ability and the best prognostic value (C-index 0.7, Figure 3a). Notably, the transcriptomic cluster-based classification outperformed the SARCULATOR nomograms in terms of prognostic value (C-index of 0.63 vs. 0.62, respectively). This suggests that, even without the incorporation of age or without being particularly designed or trained to specifically predict OS, the transcriptomic cluster-based model offers superior prognostic accuracy than the SARCULATOR nomograms (which include age as a necessary variable for its calculation). Furthermore, the TC+AGE model showed a clearly superior prognostic value than the SARC model (C-index of 0.7 vs. 0.62, respectively), which is also noteworthy.

**Figure 3.**
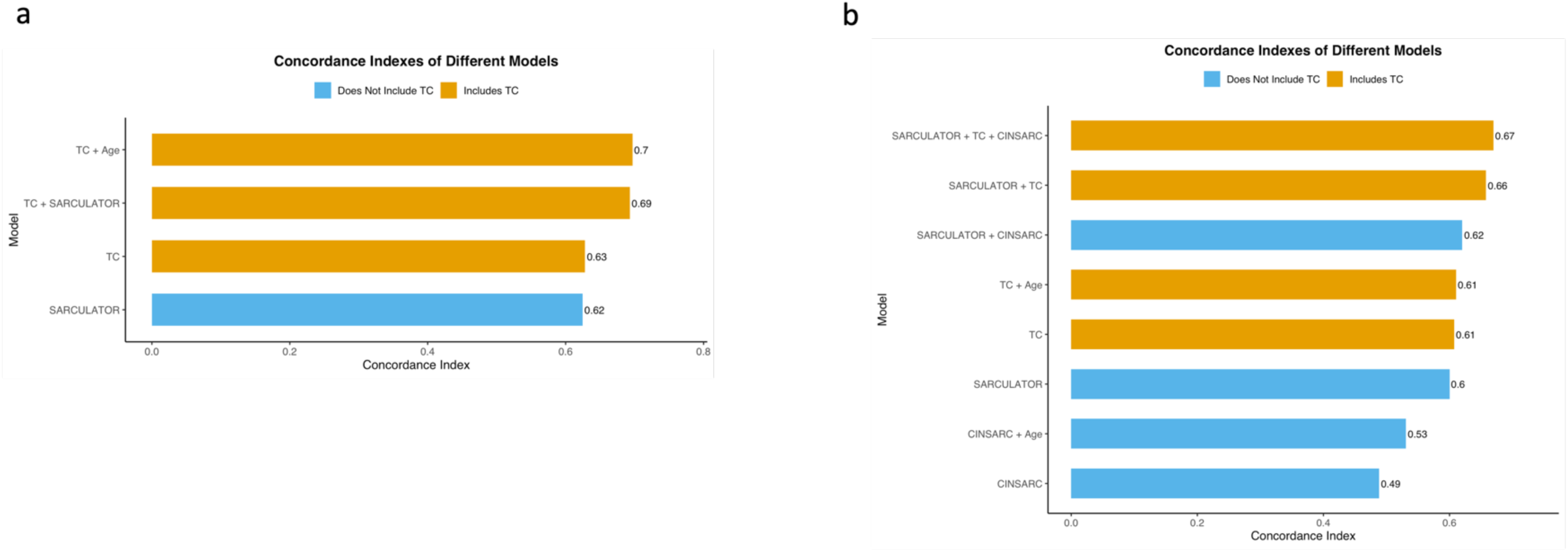
– The transcriptomic cluster-based classification outperforms the SARCULATOR clinical nomograms in terms of prognostic value. **a)** Bar chart displaying the concordance indexes of different prognostic models employed using the population of the study cohort (including SARC, TC, TC + SARC and TC + Age) **b)** Bar chart showing the concordance indexes of different prognostic models employed using the population of the validation cohort (TCGA-SARC) (including CINSARC, CINSARC + Age, SARC, TC, TC + Age, SARC + CINSARC, SARC + TC, SARC + TC + CINSARC)

Altogether, these results, in an analysis within our study cohort, point towards a superior prognostic value of the transcriptomic cluster-based classification over the currently employed gold-standard clinical nomograms approach.

### Independent validation of the superior prognostic value of the molecular signature and transcriptomic cluster-based classification

We sought to validate the superior prognostic value, when compared with clinical-based prognostication tools, of the transcriptomic cluster-based classification using an independent/external cohort. We performed the same analysis for the TCGA-SARC cohort [27], calculating the C-Indexes of different Cox Proportional Hazard Models for OS and comparing them. Distinct model combinations were considered, including SARCULATOR, CINSARC (classification of the patients of the TCGA-SARC cohort using CINSARC was possible, oppositely from what was verified for the patients of the study cohort as reported in **Results - A transcriptomic cluster-based classification outperforms the CINSARC expression-based signature in terms of prognostic value** and **Methods)** and TC as features.

In this cohort, the results of our analysis reinforced our findings, with TC outperforming, albeit marginally, SARCULATOR (C-Index of 0.61 vs. 0.6, respectively) (Figure 3b). Notably, the addition of age to the TC model did not affect its performance, with TC+AGE showing the same C-Index of 0.61 than TC.

Additionally, TC (C-Index of 0.61) consistently outperformed CINSARC (C-Index of 0.49) and CINSARC + AGE (C-Index of 0.53), demonstrating that our molecular-based classification displays a superior prognostic value than the currently used molecular-based classification.

Importantly, the best-performing models were those incorporating TC. The combination of TC with SARCULATOR achieved a C-Index of 0.66, while the combination of TC with SARCULATOR and CINSARC further improved performance, achieving the highest C-Index of 0.67. These findings validate the robustness of the transcriptomic cluster-based classification and highlight its critical role in enhancing the accuracy of prognostic models when combined with clinical and molecular predictors.

### A molecular signature and transcriptomic cluster-based classification may allow prognostic sub-stratification within specific SARCULATOR-defined prognostic groups

Additionally, we attempted to understand if the application of our transcriptomic cluster-based strategy could identify and sub-stratify patients with different prognostic horizons inside the same SARCULATOR-defined prognostic groups (predicted 5-year OS > 60% vs. predicted 5-year OS ≤ 60%).

Sub-stratification inside the favorable prognostic group, defined by patients with a predicted 5-year OS > 60%, could spot patients with a distinct prognostic profile according to the transcriptomic cluster their STS belongs to and that, in case of a relative negative prognostic profile, might benefit from an early and tailored adjuvant systemic treatment approach and/or a more intensive surveillance approach. Sub-stratification inside the unfavorable prognostic group, defined by patients with a predicted 5-year OS ≤ 60%, could identify, among the pool of patients that collectively display an indication for adjuvant chemotherapy, patients with worse relative prognosis, whose adjuvant systemic treatment approach should potentially be intensified (either in terms of number or doses of systemic treatment agents, or also in terms of frequency of treatment cycles) and patients with a better prognosis, whose adjuvant treatment approach could be, relatively, less aggressive (also in the same terms that have been previously mentioned, but in the opposite direction).

Among the 67 patients of the study cohort whose classification with SARCULATOR was amenable to be performed, 30 displayed a predicted 5-year OS > 60% and 37 showed a predicted 5-year OS ≤ 60%.

By performing a survival analysis using the Kaplan-Meier method, the transcriptomic clusters-based classification was able to significantly sub-stratify patients with different prognostic horizons within the unfavorable prognostic group (predicted 5-year OS ≤ 60%) (p-value 0.018), while it was not able to significantly sub-stratify patients with distinct prognostic profiles within the favorable prognostic group (predicted 5-year OS > 60%) (p-value 0.78) (**Supplementary Figure 6**). Considering the unfavorable prognostic group, there is a statistically significant difference in OS between patients whose STS belongs to C1 and patients whose STS belongs to non-C1 clusters (C2, C3 and C4). Patients whose STS belongs to C1 display a better relative prognosis.

Next, we performed an identical survival analysis in the TCGA-SARC cohort. The transcriptomic clusters-based classification was not able to significantly sub-stratify patients with distinct prognostic profiles either within the favorable prognostic group (predicted 5-year OS > 60%) (p-value 0.14) and also within the unfavorable prognostic group (predicted 5-year OS ≤ 60%) (p-value 0.28) (**Supplementary Figure 7a**).

If only patients from the TCGA-SARC cohort with a grade 3 DDLPS, LMS and UPS were considered, the transcriptomic clusters-based classification would still not be able to significantly sub-stratify patients with distinct prognostic profiles either within the favorable prognostic group (predicted 5-year OS > 60%) (p-value 0.32) and also within the unfavorable prognostic group (predicted 5-year OS ≤ 60%) (p-value 0.6) (**Supplementary Figure 7b**). However, it is important to note that, even lacking statistical significance, the survival curves indicates the tendency of a better prognosis for patients with a C1 STS, when compared with patients with a non-C1 STS. This apparently discordant finding (within the study cohort and within the validation cohort) may be explained by some statistical data regarding the study populations: among the 127 patients included in the TCGA-SARC that have a formal diagnosis of DDLPS, LMS and UPS, only 33 have a grade 3 STS. Of these 33 patients with a grade 3 DDLPS, LMS or UPS, only 23 display a predicted 5-year OS ≤ 60% (estimated using SARCULATOR). Our study cohort includes a higher absolute number (N=37 vs. N=23) and a higher proportion (37/70; 52,9% vs. 23/127; 18,1%) of patients with a grade 3 STS that display a predicted 5-year OS ≤ 60%.

### A transcriptomic cluster-based classification outperforms the CINSARC expression-based signature in terms of prognostic value

We also compared the prognostic values of the transcriptomic cluster-based classification with the Complexity INdex in SARComas (CINSARC) (an expression-based signature related to mitosis and chromosome integrity), using patients from the TCGA-SARC dataset with the same STS histopathological subtypes than the patients that were included in our study cohort (DDLPS, LMS and UPS) (N=127).

CINSARC annotation of our study cohort was not possible since the FoundationOne®RNA gene set does not include 32 of the genes included in CINSARC (48% of the total number of genes considered in CINSARC) (listed in detail in **Supplementary Materials)**. Theoretically, it would still be possible to classify our study cohort patient samples using CINSARC, but, conceptually, this classification would not be robust and would have a significant degree of inaccuracy.

We tested how does CINSARC overlap the lists of differentially expressed genes of each of the transcriptomic clusters. We observed an overlap of 38% between the CINSARC gene set and the C1 under expressed genes and found a significant correlation between the enrichment scores of C1_under and CINSARC (Spearman’s Rank correlation = 0.78 between NES C1 under and CINSARC (**Supplementary Figure 8**)).

Considering the TCGA-SARC patients (and after classifying them using CINSARC), an analysis using a Cox Proportional Hazards Model, including the histopathological classification (either the originally used in TCGA-SARC and the recently proposed and currently used one), CINSARC, transcriptomic clusters-based classification, and the FLNCC grade, was performed. The results revealed that the transcriptomic clusters-based classification were the only variable that showed a statistically significant correlation with OS and that the transcriptomic clusters-based classification were, subsequently, the most significant variable for the prediction of patients OS (HR 2.15; 95% CI 1.134-4.1 P=0.019) (Figure 4a). A subsequent ANOVA test of the Cox Proportional Hazards Model showed, once again, that the transcriptomic cluster-based classification was the most significant predictor of OS (Figure 4b).

**Figure 4.**
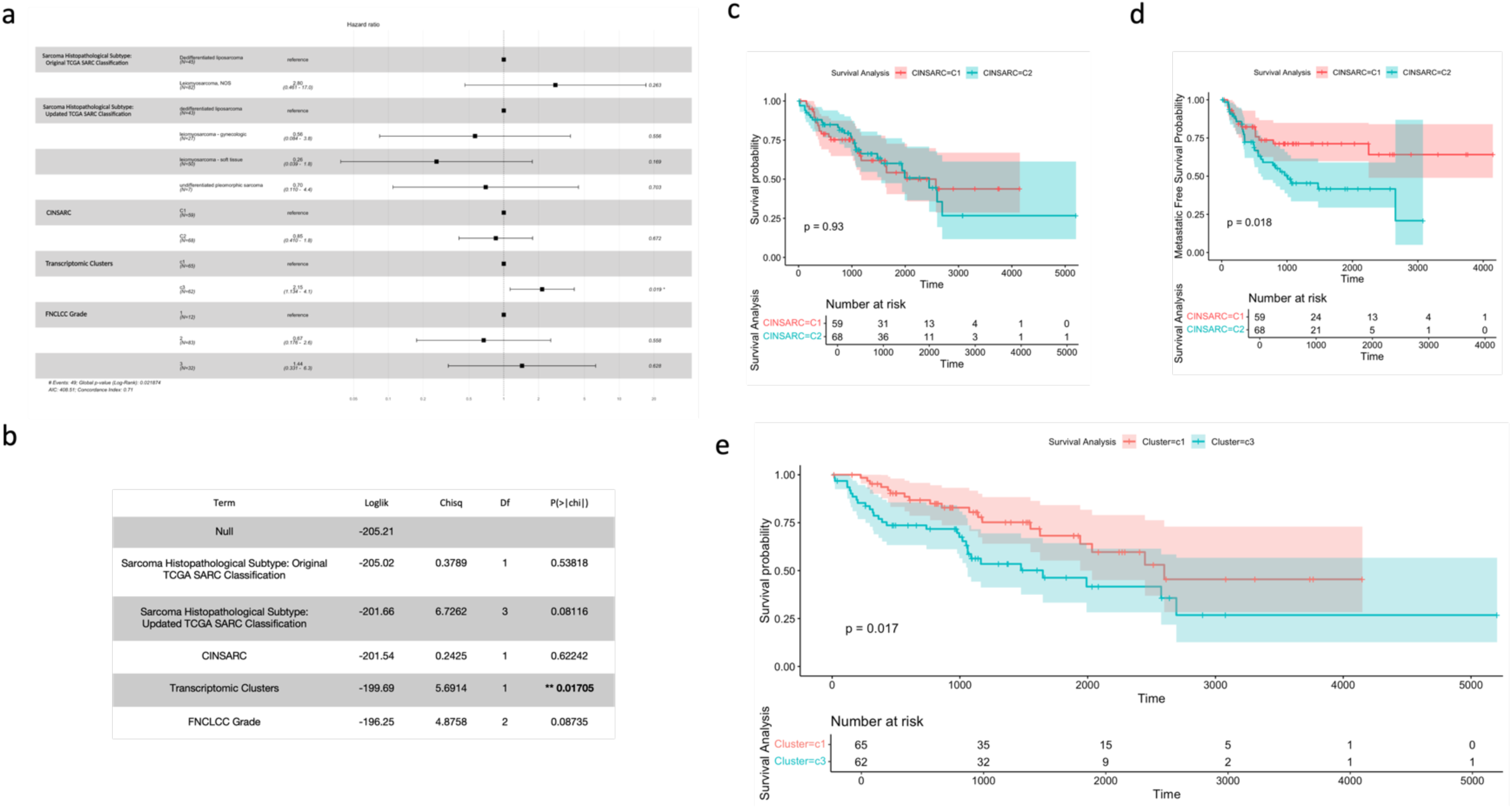
- The transcriptomic cluster-based classification outperforms the CINSARC expression-based signature in terms of prognostic value. **a)** Forest plot showing the results of the evaluation of the impact of histopathological classification, CINSARC, transcriptomic clusters-based classification, and the FLNCC grade on OS considering the patients of the TCGA-SARC cohort after their classification accordingly to CINSARC. **b)** Table displaying the results of the ANOVA test applied to the Cox Proportional Hazards Model to assess the predictive ability of different variables for OS estimation using data from the TCGA-SARC (after the classification of TCGA-SARC patients accordingly to CINSARC). **c)** Survival analysis of the patients of the TCGA-SARC cohort after their classification accordingly to CINSARC: OS analysis by the Kaplan-Meier method and respective curves (time scale is shown in days). **d)** Survival analysis of the patients of the TCGA-SARC cohort after their classification accordingly to CINSARC: MFS analysis by the Kaplan-Meier method and respective curves (time scale is shown in days). **e)** Survival analysis of the patients of the TCGA-SARC cohort after their classification in accordance with the transcriptomic clusters-based classification: OS analysis by the Kaplan-Meier method and respective curves (time scale is shown in days).

Additionally, an OS analysis by the Kaplan-Meier log rank test was performed for the CINSARC classified patients (C1 and C2) and showed no statistically significant differences (log rank P=0.93) (Figure 4c). On the other hand, CINSARC has the ability, as previously reported, to distinguish between C1 and C2 in terms of metastasis free survival (MFS), displaying a log rank of P=0.018 (Figure 4d).

In parallel, TCGA-SARC patients were also classified according to the transcriptomic clusters-based classification (as previously mentioned) and a survival analysis employing the Kaplan-Meier method showed, in this case, a significant difference in OS between C1 and C3 (Log Rank P=0.01) (Figure 4e).

These findings confirm that, despite being able to accurately predict MFS, CINSARC does not have the capacity to differentiate distinct OS profiles within STS patients and displays a lower overall prognostic value than the transcriptomic clusters-based classification.

### The analysis of the DNA alterations (detected with FoundationOne®CDx) found in the patients included in each of the 4 transcriptomic clusters reveals unique actionable targets

We analyzed the DNA alterations detected by FoundationOne®CDx in the patients included in each of the 4 transcriptomic clusters (**see Methods**). The frequency and types of the detected genomic alterations are represented in Figure 5a. An extensive description of these genomic alterations, and their respective distribution per transcriptomic cluster, is provided in **Supplementary Material**.

**Figure 5.**
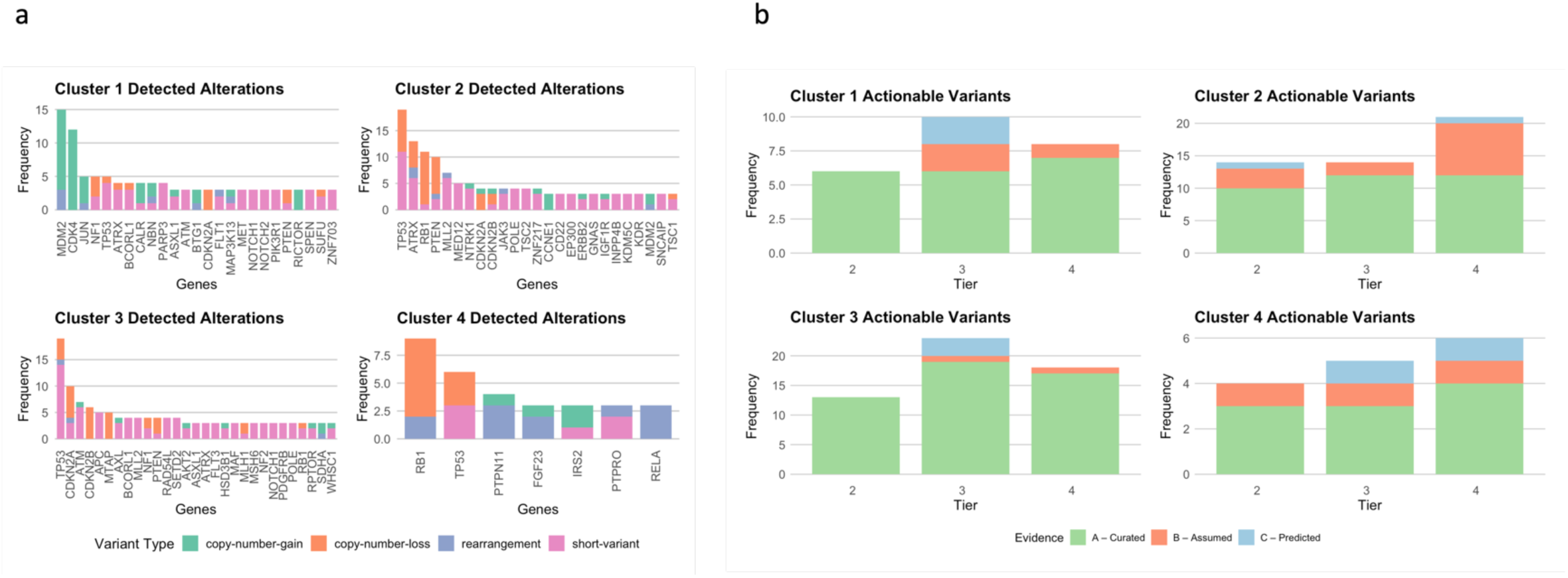
– The analysis of the DNA alterations detected with FoundationOne®CDx for patients included in each of the 4 transcriptomic clusters reveals unique actionable targets. **a)** Frequency and types of genomic alterations detected by FoundationOne®CDx for patients included in each of the transcriptomic clusters. **b)** Distribution of the detected genomic alterations classified with MTBP per tier of actionability (Tier 2 – Investigational, Tier 3 – Hypothetical Target: Alteration-drug match is associated with antitumor activity, but magnitude of benefit is unknown (potential cancer-repurposing opportunity, Tier 4 – Hypothetical Target: preclinical evidence of actionability) and per functional relevance evidence for the alteration (A – Curated; B – Assumed; C – Predicted) for each of the transcriptomic clusters.

Then, we used MTBP [28] to systematize and interpret the functional and predictive value of each of the genomic variants that were found for the patients included in each of the transcriptomic clusters (**Supplementary Tables 3,4,5** and **6**). The functional classification and the actionability tiering for gene variants performed by MTBP follow the ESMO Scale of Clinical Actionability for Molecular Targets (ESCAT). The distribution of the detected genomic alterations per tier of actionability and level of evidence for each of the transcriptomic clusters is shown in Figure 5b.

Overall, 151 gene variants classified with ESCAT evidence tiers ranging from 2 to 4 have been identified among the patients included in the study cohort (29 clinically actionable gene variants have been identified for patients included in C1, 51 for patients included in C2, 56 for patients included in C3, and 15 for patients included in C4). C2 displayed the highest number of gene variants classified with an ESCAT evidence tier 2 (15 variants), followed by C3 (14 variants). C3 showed the most significant number of gene variants classified either with an ESCAT evidence tier 3 (24 variants) and with an ESCAT evidence tier 4 (18 variants). C4 presented the lowest number of gene variants classified with an ESCAT evidence tier 2 (4 variants), with an ESCAT evidence tier 3 (6 variants) and with an ESCAT evidence tier 4 (5 variants).

**Supplementary tables 3, 4, 5 and 6** display, extensively and in full detail, the particular features of each of the gene variants that were found and their distribution per transcriptomic cluster and per ESCAT evidence tier.

Among the complete pool of gene variants classified with an ESCAT evidence tier 2, there is a ubiquitous presence (across all the 4 clusters) of *MDM2* amplifications (conferring sensitivity to Brigimadlin and Milademetan) and a vast plethora of *TP53* alterations (mainly missense mutations conferring sensitivity to Pazopanib and Vorinostat). *MTAP* deletions (conferring sensitivity to MRTX1719 and AMG193) are also noteworthy, since they were found in 3 of the 4 clusters. *TSC2* mutations (conferring sensitivity to ABI-009) were also identified in 2 of the 4 clusters. Interestingly, *ERBB2* amplifications (conferring sensitivity to Trastuzumab Deruxtecan) (found in C2) and *PIK3CA* missense mutations (conferring sensitivity to Capivasertib and Copanlisib) (found in C3) were found in 1 of the 4 clusters.

C1 is marked, in terms of actionable alterations, by an enrichment in *MDM2* amplifications (Tier 2), *TP53* mutations (Tier 2), *NF1* mutations (Tier 3 – conferring sensitivity to Selumetinib and resistance to Vemurafenib; - and Tier 4 – conferring sensitivity to Trametinib and Cobimetinib) *CDK4* amplifications (Tier 4), and alterations of different genes involved in HRR, namely frameshift mutations of *RAD51B* (Tier 3), and missense mutations of *ATM* (Tier 3) and *BRIP1* (Tier 3), all of them conferring sensitivity to PARP inhibitors, namely Olaparib.

C2 is characterized, besides *MDM2* amplifications (Tier 2) and *TP53* mutations (Tier 2), by *MTAP* deletions (Tier 2), *TSC2* mutations (Tier 2), *ERBB2* amplifications (Tier 2 – conferring sensitivity to Trastuzumab Deruxtecan – and Tier 3 – conferring sensitivity to a wide array of anti-HER2 agents, either in monotherapy or in combination with other drugs belonging either to the same anti-HER2 class or to other classes) and specific genetic alterations that are targets for tumor agnostic treatment approaches, such as *RET* missense mutations (Tier 3, which confer sensitivity to Selpercatinib and Pralsetinib). *POLE* missense mutations (Tier 3), that typically lead to an hypermutated and immunosensitive phenotype, conferring sensitivity to immune-checkpoint inhibitors such as Pembrolizumab, and *FGFR1* mutations (Tier 3 – conferring sensitivity to Pemigatinib – and Tier 4 – conferring sensitivity to Erdafitinib and AZD4547) are also of note.

C3 is portrayed by *MDM2* amplifications, *TP53* mutations, *MTAP* deletions, *TSC2* mutations, and *PIK3CA* mutations in terms of Tier 2 alterations. This cluster is particular fertile in terms of actionable alterations. A mention should be made to *POLE* missense mutations (Tier 3, conferring sensitivity to Pembrolizumab), *KRAS* missense mutations (Tier 3 and Tier 4, conferring sensitivity and resistance to a plethora of different agents), *NRAS* missense mutations (Tier 3 and Tier 4, conferring sensitivity and resistance to a plethora of different agents), *MET* amplifications (Tier 3, conferring sensitivity to Capmatinib, Tepotinib, Telisotuzumab Vedotin and Crizotinib), *PTEN* frameshift mutations (Tier 3 – conferring sensitivity to Capivasertib and Fulvestrant – and Tier 4 – conferring sensitivity to Ipatasertib, GSK26364771, AZD8186), *VHL* missense mutations (Tier 3 – conferring sensitivity to Everolimus), *CDK4* amplifications (Tier 4 – conferring sensitivity to cyclin-dependent kinases inhibitors), *CDKN2A* mutations (Tier 4 – conferring sensitivity to cyclin-dependent kinases inhibitors), and alterations of genes involved in the HRR mechanisms, namely *ATM* (Tier 3), and *ATR* (Tier 3), conferring sensitivity to PARP inhibitors.

C4 is the cluster with the smallest number of actionable gene alterations among the 4 clusters. *MDM2* amplifications (Tiers 2, 3 and 4), TP53 mutations (Tiers 2, 3 and 4), *MLH1* missense mutations (Tier 3, conferring sensitivity to PARP inhibitors), *BARD1* missense mutations (Tier 3, conferring sensitivity to PARP inhibitors) and *CDK4* amplifications (Tier 4, conferring sensitivity to cyclin-dependent kinases inhibitors) are found in this cluster.

Overall, there is a profusion of targetable alterations scattered across the different transcriptomic clusters. Either alterations that are compelling targets for tumor-agnostic treatment approaches, such as the *RET* mutations and *ERBB2* amplifications found in C2, or alterations that are linked with defective DNA repair mechanisms, such as the mutations of distinct genes involved in HRR verified in C1, C3 and C4 and the mutations of *POLE* documented in C2 and C3, and a vast array of other specific alterations (some of them never previously documented in DDLPS, LMS and UPS) that confer sensitivity to a broad spectrum of different agents.

### RNAseq detected fusions that were not identified by DNAseq

FoundationOne®RNA detected fusions that were not detected by FoundationOne®CDx in 9.1% of the cases (2/22) for which both DNAseq and RNAseq for rearrangement detection were clinically reportable. The inclusion of a high number of archival samples > 2 years old contributed to the high rate of absence of passage of the particularly rigorous post-sequencing QC metrics required for clinical RNA rearrangement detection. On the other hand, the great majority of samples (75/102; 73.5%) passed the QC metrics required for RNAseq expression analysis (see **Methods**). The disparity between these QC passage rates for the optimal detection of cancer-related gene fusions and rearrangements (318 gene panel) test and the gene expression profiling (1517 gene panel) test covered by FoundationOne®RNA lies on the different nature of each of these tests: the test designed for optimal detection of cancer-related gene fusions and rearrangements for 318 genes is a test developed for clinical use and, therefore, employs specially stringent QC criteria, while the gene expression profiling test for 1517 genes is a test designed for research use only and employs less strict QC criteria (see **Methods**). The STS histotypes that were provided for this study are not typically translocation-associated types, so the limited number of detected fusions is relatively unsurprising. An *HMGA2* (intron 3)::*TPH2* (intron 8) fusion was found in a case of DDLPS. This fusion was not detected in DNA because the *HMGA2* and *TPH2* genes are not baited on the FoundationOne®CDx gene panel. A *NOTCH3* (intron 24)::*BRD4* (intron 11) fusion was found in a case of UPS. Similarly, this fusion was not detected by FoundationOne®CDx because, while the exonic regions of both genes are covered on FoundationOne®CDx, the breakpoints for both genes occurred in intronic regions which are not covered. Thus, RNAseq provided additional value to DNAseq by detecting reportable fusions.

## Discussion

In this study, the analysis of RNAseq data from a cohort composed by 102 samples of the most common STS subtypes using unsupervised machine learning models allowed the unravel of previously unknown molecular patterns and permitted the identification of 4 well-defined transcriptomic clusters. These transcriptomic clusters have a clear prognostic value, which was externally validated. The prognostic value of this transcriptomic cluster-based classification seems to be superior to currently used clinical-based prognostication tools (such as SARCULATOR nomograms) and to modern gold-standard molecular-based prognostication tools (such as CINSARC). The analysis of DNAseq data from the same cohort of STS samples revealed a plethora of unique and, in some cases, never documented molecular targets for precision treatment across different transcriptomic clusters.

Clustering, the concept of grouping samples/patients based on the co-occurrence of molecular alterations, has been previously used to systematically analyze the significant amounts of complex data generated by bone and soft tissue sarcoma molecular characterization approaches (either single or multi-omics), allowing the identification of specific sarcoma molecular clusters with particular clinical behaviors [27; 29-32]. Consensus clustering, the unsupervised machine learning model that we employed in this study, has also already been used in two studies to identify STS molecular clusters [20,33] (**Supplementary Table 7**). Besides differences in terms of representation of distinct STS histopathological subtypes in sample pools, molecular profiling approaches, methodological strategies for data analysis (combination with other types of unsupervised clustering or with different methods) and respective results, the distinctive feature of our approach is its clinical-driven nature. Our method has been primarily developed using analytical tools developed for research use, but subsequently powered with analytical tools with a proven clinical utility and has included an extensive gathering of a wide array of clinical variables, allowing a better portrayal of the clinical significance of the clusters and its defining molecular features. We used sequencing tests that have been developed for research and/or clinical use, are cost-effective and are, therefore, potentially useful in the clinical practice routine. These tests were used to analyze all the samples of the study cohort, solely for the purpose of this study, differing from an approach comprising the analysis of a previously constructed public database. Ultimately, we have identified gene expression signatures that display both a superlative prognostic and also a potential predictive value, supreme indicators of clinical significance and impact.

The conceptual robustness of the identified transcriptomic clusters is supported by the methodological approach (use of the Elbow method and verification of clusters persistence with the removal of UPS samples from the samples pool), the presence of all of the included STS histopathological subtypes in each of the clusters and the distinct intrinsic nature of the molecular features that define each cluster.

Various of these cluster defining molecular traits are, themselves, novel and constitute, in some cases, breakthrough findings in STS (**Supplementary Table 8** [20; 34-48]). Some of these particular molecular characteristics have never been previously reported in STS. The over expression of specific MAGE genes (*-A12, -A2B, -A3, -B1, -B2*, and - *C2*), other than *MAGE-A4* [36,37], verified in C2, the over expression of HLA class II (*HLA-DMA, HLA-DMB, HLA-DOA, HLA-DQA, HLA-DRA* and *HLA-DRB1*) genes, other than HLA class I genes [41,42], verified in C3 and the over expression of claudin 4 verified in C4 fall into this category.

Other cluster defining molecular alterations have already been reported in STS as exceedingly rare findings. The under expression of several genes involved in HRR mechanisms, potentially leading to homologous recombination deficiency (HRD), in C1 is an example [27,34]. The over expression of *SSX* genes (*−1, −2, −2B* and *−3*) documented in C2 is another illustrative case, especially considering that the sample pool did not include synovial sarcoma samples (even though the over expression of *SSX* genes may also be found in other STS subtypes, with a significant fraction of these STS’s co-over expressing more than one *SSX* family member [37–39]).

From another angle, the coexistence of some of these cluster-specific molecular traits hasn’t also been previously described in STS. The coexistence of over expression of *CDK4* and under expression of genes involved in HRR, as verified in C1, is exemplifying. Even though the concomitant over expression of *MAGE* and *SSX* genes has already been reported in colorectal cancer (being correlated with the development of metastasis to the liver [40]), the simultaneous over expression of the prementioned specific *MAGE* genes and the aforesaid *SSX* genes, as verified in C2, has never been reported in the STS histotypes that compose our cohort. **Supplementary Table 8** lists distinctive molecular traits per cluster, and a conceptual framing of its rarity or novelty based on a literature review.

Besides the originality of the molecular features that are the backbone of each cluster, the biological and clinical relevance of the transcriptomic clusters also lie in their superlative prognostic value. Three of the four identified clusters within our cohort have a clear prognostic value, and the associated molecular signatures show, when compared with the histopathological classification and other variables, a better ability to predict OS, a finding that was externally validated with the TCGA-SARC cohort. This embodies the notion that a molecular-based classification captures the biological behavior and estimates prognosis more accurately than the conventional histopathological classification.

Given the intrinsic prognostication power of the clusters, it is crucial to understand if the exactness of prognosis estimation that they offer is superior to the prognostication precision offered by clinical (SARCULATOR nomograms) and molecular-based (CINSARC) tools that are currently considered gold-standard and widely used in clinical practice.

Our transcriptomic cluster-based classification, which was not originally designed or trained to specifically predict OS, exceeded the SARCULATOR nomograms prognostic value and, when combined with age, displayed the strongest OS predictive ability among different individual variables (including SARCULATOR) and variables combinations in an analysis within the study cohort. This was independently validated using the TCGA-SARC cohort, even considering the marginal difference verified between the prognostic accuracy of the transcriptomic clusters-based classification and SARCULATOR.

Additionally, the present study cohort includes, up to a certain point, STS populations that were underrepresented in the cohorts used for the development of the past mentioned nomograms, including eSTS, RPS and trunk STS concomitantly, a small number of patients treated in a neoadjuvant context (3/101) and a small number of patients with an unresectable RPS (2/101). Furthermore, our results show that the use of molecular data, that may be obtainable from the sequencing of a biopsy specimen, or the combination of a variable that is objective and independent from a histopathological examination, age, and molecular data, is superlative, making this prognosis-estimation strategy potentially employable in a preoperative setting, oppositely from the available nomograms as some of the variables included for their calculation are not available before surgery [49]. Finally, our molecular-based classification may refine SARCULATOR’s prognosis assessment, allowing prognostic sub-stratification within specific SARCULATOR-defined prognostic groups following the analysis of the study cohort. This was not verified when the TCGA-SARC cohort was used for validation potentially because of the different preponderance, in comparison with the study cohort, of patients with a grade 3 STS that display a predicted 5-year OS ≤ 60% in this cohort, as reported in **Results.**

Our transcriptomic clusters-based classification is, as CINSARC, an expression-based signature established from the analysis of primary non-translocation-related STS [25,50]. A comparative analysis of the prognostic value of two STS molecular-based signatures whose prognostic value outperformed the histopathological-based grading system in STS is therefore essential. The overlap between the CINSARC gene set and the C1 under expressed genes and the significant correlation between the enrichment scores of C1_under and CINSARC is not surprising considering the molecular features that characterize CINSARC expression (the 67 genes contemplated in CINSARC are involved in the control of chromosome integrity and mitosis, and CINSARC expression is associated with genomic and chromosomal instability [25,50]) and the molecular features that characterize C1_under (under expression of genes involved in HRR, potentially leading to HRD and chromosomal instability).

Methodologically, we employ ssGSEA to derive pathway enrichment scores from normalized gene expression data, encompassing both KEGG pathways and our custom gene sets. Following this, we extract p-values for each pathway and apply an adjustment as mentioned in **Methods**. This adjustment is performed both across patients and pathways, ensuring that the comparisons are robust and reducing the likelihood of false positives. After excluding KEGG pathways, samples are assigned to the pathway with the minimal adjusted p-value. In contrast, CINSARC classifies samples by calculating the distance between a sample’s normalized gene expression data and the centroids of CINSARC gene set. Samples are then classified to the closest centroid with some samples remaining unclassified if the threshold is not met. While the CINSARC approach relies on the proximity of a sample’s gene expression to the nearest centroid within predefined gene sets (CINSARC C1 and C2), our method is based on adjusted ssGSEA enrichment scores. By incorporating KEGG pathways and adjusting p-values both across patients and pathways, our method provides a robust strategy of representing the enrichment, which is adjusted for pathway and sample-wise false positives.

Apart from the methodological dimension, the nature of the clinical endpoints that may accurately be estimated following the use of CINSARC or our transcriptomic cluster-based method is also distinct. The OS estimation capacity of our transcriptomic cluster-based clearly surpasses CINSARC in a head-to-head comparison using the TCGA-SARC cohort, as shown in **Results**. Our analysis also show that, using the same cohort, while CINSARC accurately differentiates patients with different MFS profiles, it does not have the power to discriminate groups of patients with different OS profiles, something that our transcriptomic cluster-based approach is capable of. While CINSARC accurately predicts MFS, our approach more precisely predicts OS.

Past series, either in a real-world context (analysis of the clinical impact of comprehensive genomic profiling and subsequent discussion of management in an institution’s molecular tumor board (MTB)) [18] or in an investigational context (analysis of the effect of the enrollment in biomarker-matched early-phase clinical trials on clinical outcomes) [51] have demonstrated that a significant percentage of STS display druggable molecular alterations and that both STS patients treated using a molecular-guided personalized treatment in a conventional context [18] and STS patients enrolled in biomarker-matched early-phase clinical trials [51] show a significant benefit from the employment of a molecular-guided strategy.

The analysis of DNAseq data of patients included in each transcriptomic cluster highlighted 151 actionable gene variants, comprising either alterations that are putative targets for tumor-agnostic treatments and also alterations that are targets for tumor-specific approaches, conferring sensitivity to a variety of molecularly targeted agents and new antineoplastic drug classes. While the most broadly represented genes for which alterations were found across different clusters overlap genes for which alterations have more commonly been reported in other series (i.e. *TP53, MDM2* and *PIK3CA*) [18], we managed to identify alterations and targets whose existence in STS has been under reported or never documented (i.e. *ERBB2* amplification, *MET* amplification, *POLE* mutations, *RET* mutations, *KRAS* and *NRAS* mutations). Notably, RNAseq identified two fusions not detected using DNAseq (*HMGA2*::*TPH2* in a case of DDLPS and *NOTCH3*::*BRD4* in a case of UPS) despite RNAseq QC metrics being sufficient for rearrangement detection in only a small percentage of samples (n=22).

While the great majority of patients included in past STS molecular-profiling series were patients in an advanced setting (most of them heavily pretreated in a metastatic context), the great majority of the patients (95%) included in our series presented with localized disease. Nevertheless, real-world series also included patients without metastases at the time of their case discussion in an MTB (15%) [18], having recommended the addition of molecularly targeted agents to a conventional chemotherapy backbone in some of the patients with actionable alterations and an early-stage disease setting (2 out of 10 patients) [18] and used, as we did in our series, primary tumor samples for molecular profiling (50% of included patients) [18] with similar conceptual results. Biologically, a recent groundbreaking study characterized the genomic differences between early-stage untreated primary tumors and late-stage treated metastatic tumors [52]. This study included primary and metastatic samples of LMS (15 vs 47) and liposarcoma (17 vs 25) [52]. No significant variations in clonality, karyotype, mutational burden, mutational signature profile, total number of driver gene alterations, frequency of therapeutically actionable gene variants and treatment-associated driver genes were found between primary and metastatic samples of both LMS and liposarcoma, but one cannot ignore that exposure to treatment (either chemotherapy or radiotherapy) potentially further scars the tumor genome and introduces an evolutionary bottleneck that may select for therapy-resistant drivers [52], making molecular profiling of metastatic lesions in an advanced setting recommendable.

Although the methodology and the findings of our work are, as shown previously, robust, there are some limitations that are important to underline. The retrospective nature of the study should be taken into account. This study cohort is single-centered (even though it includes patients with different ethnical ancestries and backgrounds – European and African-native patients). Besides this, this study population and samples pool is comprised by a limited number of 3 STS histopathological subtypes, and is composed by primary tumor samples of a great majority (95%) of STS patients with early stage/localized disease. Therefore, there is not a significant representation either of samples from STS metastases, and also of patients with advanced STS. Moreover, the samples included in each of the batches that were sent to Foundation Medicine for DNA and RNA sequencing were collected in different timepoints and, therefore, display heterogenous chronological ages (a fact that directly impacts the differential likeability of degradation of the biological material and the distinct quality of the samples for the planned sequencing analysis). In the same line, the degree of degradation of the samples that were analyzed and, subsequently, the amount of samples for which the quality control for DNA and RNA sequencing was not successful, namely in the in the context of fusion/splice site detection with RNA, is also a limitation.

On the other hand, the sequencing tests that were used for molecular profiling are targeted sequencing tests, which offer results with a distinct conceptual coverage than the ones that could be offered by a whole genome or whole exome sequencing approach.

Although the gene set of this targeted sequencing test (namely Foundation One^®^ RNA) has not been primarily created specifically using a particular panel of genes whose differential expression profile portrays and is characteristic of STS, it covers 52% of the genes that comprise the gene set of the single molecular signature that is based on specific gene expression profiling in STS (CINSARC), shows an overlap of 38% between the pattern of expression of a specific array of covered genes and the pattern of expression of the genes that compose CINSARC’s gene set (with a Spearman’s rank correlation of 0.78), displays a provenly superior OS predictive capacity and prognostic value when compared with CINSARC (a finding that has been externally validated using the TCGA-SARC dataset), and the subsequent newly found transcriptomic cluster-based classification is possibly the first molecular-based classification that has the capability of predicting OS in STS.

## Methods

A detailed description of the specific contribution of each of the participating institutions - Instituto de Medicina Molecular João Lobo Antunes (iMM), Instituto Português de Oncologia de Lisboa Francisco Gentil (IPOLFG), Instituto Superior Técnico (IST), F.Hoffmann-LaRoche and Foundation Medicine - may be found in Author Information - Contributions (see **Contributions**).

Ethical considerations are also provided in Ethics Declarations (see **Ethics Declarations**).

### Sample characterization

This study has included 102 formalin-fixed paraffin-embedded (FFPE) neoplastic tissue samples from 101 STS patients diagnosed and treated at IPOLFG (a tertiary oncological center, one of the sarcoma European reference centers) between the 15^th^ of April 2013 and the 29^th^ of September 2022.

These samples were previously stored at the IPOLFG tumor biobank, and were part of this biobank sarcoma collection.

The sample pool was comprised by 26 DDLPS samples, 25 high-grade LMS samples and 51 UPS samples.

A sarcoma-dedicated pathologist reviewed each of the 102 STS samples. The pathologist scored the images for all the 102 samples that were shipped to Foundation Medicine, Inc. (Cambridge, Massachusetts, United States of America) for molecular profiling. The number of slides available for review from each case ranged from 1 to 6. Pathology reports were reviewed for sarcoma site, depth, FNCLCC grade, presence of multifocality, completeness of resection, reported immunohistochemical studies and/or molecular diagnostics and, subsequently, histopathological diagnoses.

The research team analyzed clinical files, retrieving data not only from IPOLFG institutional records, but also from accessible national electronic clinical files. An anonymized database has been developed specifically for this study. This database includes detailed information on patient demographics, sarcomas’ characteristics, treatment strategies (both neoadjuvant and adjuvant), surgical data, and oncological follow-up. The most recent follow-up has been conducted on October 10^th^, 2023.

### Samples circuit

A formal histopathological review was firstly conducted at IPOLFG. The FFPE blocks were then transported to iMM where they were sectioned by the Comparative Pathology Unit team. The slides obtained were stored at the Translational Oncobiology Laboratory at iMM, while the blocks were shipped to Foundation Medicine, Inc. The samples were shipped in three different batches – the first one, including 26 samples of DDLPS, was shipped on 07/2022; the second one, including 25 samples of LMS, was shipped on 10/2022; the third one, including 51 samples of UPS, was shipped on 01/2023.

### DNA and RNA sequencing

102 FFPE STS samples from 101 patients were characterized using FoundationOne^®^CDx (F1CDx^®^) for DNA sequencing (DNAseq) and FoundationOne^®^RNA (F1RNA) for RNA sequencing (RNAseq). Testing was performed in a Clinical Laboratory Improvement Amendments (CLIA)-certified, College of American Pathologists (CAP)-accredited, New York State-approved laboratory (Foundation Medicine, Inc., Cambridge, MA, USA). DNA and RNA were simultaneously co-extracted and isolated from FFPE samples. F1CDx is a next generation sequencing (NGS)-based assay for the detection of short variants (substitutions and short insertions/deletions [indels]), copy number alterations (CNAs), and large genomic rearrangements in 324 cancer-associated genes, as well as reporting of complex biomarkers including microsatellite instability (MSI) and tumor mutational burden (TMB). The clinical and analytical validation for F1CDx has been published by Milbury et al. [53]. F1RNA is a laboratory developed test that uses hybrid-capture based targeted RNAseq designed for optimal detection of cancer-related gene fusions and rearrangements for 318 genes for clinical use and gene expression profiling (GEP) for 1517 genes for research use only (RUO). Analytical validation studies for fusion detection have been previously performed to assess fusion calling accuracy, reproducibility, and limit of detection in 189 clinical solid tumor specimens [54]. The results from both DNAseq and RNAseq were periodically sent back to iMM, IPOLFG, and IST via an encrypted and safe platform.

### DNAseq and RNAseq data analysis

#### Population considered for molecular analysis

Out of 102 STS samples sent for molecular analysis, 79 samples passed F1CDx quality control and were sufficient for DNA analysis, and 75 samples passed F1RNA quality control and were sufficient for RNAseq expression analysis. One additional sample was excluded from the RNAseq expression analysis after being identified as an outlier using principal component analysis (PCA). A total of 74 samples (16 DDLPS, 15 LMS and 43 UPS) were therefore considered for downstream expression analysis.

Of note, 53 of the 75 samples were excluded for analysis of fusions in RNA due to not passing post-sequencing QC metrics required for clinical RNA rearrangement detection. As mentioned in **Results**, the disparity between these QC passage rates for the optimal detection of cancer-related gene fusions and rearrangements (318 gene panel) test and the gene expression profiling (1517 gene panel) test covered by FoundationOne®RNA lies on the different stringentness of each of these tests, considering that one has been developed for clinical use and other has been designed for research use only.

#### RNAseq expression data analysis - Transcriptomic clusters discovery

The computational analyses were performed using R (v4.4.0). RNAseq data from these 74 samples was filtered for expression using the edgeR (v4.2.1) [55] filterByExpression method to remove lowly expressed genes, followed by Voom normalization to stabilize variance across samples. The genes were then filtered based on Mean Absolute Deviation, retaining the top 55% of the most variable genes. Consensus Clustering from the ConsensusClusterPlus package (v1.68.0) [56] was applied and evaluated using the Elbow Method, which identified an optimal number of 4 clusters. Differential gene expression analysis was then conducted through pairwise comparisons between clusters using the limma package (v3.60.4) [57] and p-values were adjusted for multiple hypothesis testing using Benjamini-Hochberg False Discovery Rate (BH-FDR). The unique genes defining each cluster were identified by intersecting the genes that were differentially expressed in the same direction within a specific cluster.

#### DNAseq genomic alteration data analysis – Genomic alterations/variants (found in the patients included in each transcriptomic cluster) actionability evaluation

We extracted the alterations detected by F1CDx in the patients included in each of the 4 transcriptomic clusters. To systematically analyze the actionability of each of the alterations highlighted, the Karolinska Molecular Tumor Board Portal (MTBP) [28] was used. MTBP offers a general framework for the interpretation of the functional and predictive value of a given list of cancer genomic variants by using several computational tools and databases that are referenced in the provided results.

### Transcriptomic clusters clinical significance assessment

#### Evaluation of transcriptomic clusters intrinsic prognostic value (study cohort)

An overall survival (OS) analysis was performed using survival package (v3.7-0) [58]. This analysis only contemplated patients whose samples were considered for the RNAseq expression analysis. 74 patients (corresponding to 74 samples) were integrated. Out of these 74 patients, 4 had not been submitted to a surgical approach and were, therefore, excluded. In total, 70 patients were considered for this analysis. Each sample was accordingly classified using Consensus Clustering as described in “RNAseq data analysis – Transcriptomic clusters discovery”. The distribution of these samples per transcriptomic cluster was considered for OS estimation. A Cox Proportional Hazards Model for OS was then applied, also incorporating other relevant clinical variables. The Schoenfeld residuals were used to validate each variable, ensuring time independence. As a result, distant metastasis was excluded from the model due to its violation of this assumption. An Analysis of Variance (ANOVA) test was subsequently performed on the Cox Proportional Hazards Model.

The TCGA-SARC dataset was used as a validation dataset [27]. This dataset was first filtered for specific subtypes (DDLPS, LMS, UPS) (N=127). It was processed similarly to our dataset, i.e. normalized using the edgeR (v4.2.1) filterByExpression method followed by Voom quantile normalization from the Limma package. After normalization, single-sample gene set enrichment analysis was performed using the corto package (v1.2.4) [59], with KEGG pathways (N=186) and the gene sets corresponding to under- and over-expressed genes in each cluster (C1_under, C1_over, C2_under, C2_over, C3_under, C3_over, C4_under, C4_over) passed as inputs. P-values for each pathway were normalized using BH-FDR and KEGG pathways were subsequently filtered out after adjusting p-values across the entire set of pathways. By considering KEGG pathways in our enrichment analysis, we were able to assess the enrichment of each of the transcriptomic clusters in terms of specific pathways and compare it with other biological processes. Finally, each TCGA-SARC sample was classified based on the gene set with the most significant p-value (P < 0.05). Similar to previous OS analysis, the Kaplan-Meier log-rank test for OS was then applied to compare the survival outcomes across clusters, and the Cox Proportional Hazards Model for OS was again employed, incorporating relevant clinical variables.

#### Exclusion of a potential contamination effect by the preponderance of UPS samples in the global pool of samples used for this analysis and in the composition of the majority of the transcriptomic clusters: focus on the persistence of patterns of molecular enrichment and prognostic value of transcriptomic clusters

Considering that the most commonly represented STS histopathological subtype in 3 of the 4 identified transcriptomic clusters is UPS and the pronounced relative preponderance of UPS in transcriptomic clusters 3 and 4, a potential “contamination” effect by UPS samples and the notion that these 4 clusters could be portraying solely the UPS molecular landscape and respective intrinsic subtypes had to be ruled out. Focusing our attention on the study cohort, the removal of the UPS samples would lead to a number of remaining samples (DDLPS and LMS) that would be too low to allow the performance of an unsupervised consensus clustering analysis. Therefore, using the TCGA-SARC dataset, we evaluated if the removal of UPS patients from the patients pool would alter the previously verified molecular enrichment of these STS patients samples in the transcriptomic clusters and would modify any statistically significant correlation that had been previously verified between the transcriptomic clusters-based classification and OS.

#### Evaluation of transcriptomic clusters relative and comparative prognostic value (external cohorts)

Patients of the study cohort were classified, using the specific clinical nomograms (either for RPS and eSTS) available at SARCULATOR (https://www.sarculator.com/), to estimate the 5-year survival probability for each patient. Then, they were stratified according to SARCULATOR’s predefined prognostic groups (5-year OS >60% vs. 5-year OS ≤60%). Out of the 70 patients whose samples were considered for the RNAseq analysis and that were submitted to a surgical approach, 67 were successfully classified using the up mentioned nomograms. The patients for whom the nomograms could not be applied, were not classified either due to lack of crucial data necessary to use the nomograms or due to the presence of tumor fragmentation, which prevented accurate estimation of tumor size. Various C-index values were then compared, derived from the Cox Proportional Hazards Model for OS. The comparisons included the following models: SARCULATOR 5-year OS prediction; transcriptomic clusters; SARCULATOR 5-year OS prediction combined with transcriptomic clusters; and finally, transcriptomic clusters combined with age. Taking the SARCULATOR stratified patients (5-year OS >60% vs. 5-year OS ≤60%), Kaplan-Meier curves were generated for each of the transcriptomic clusters and a comparative analysis was then performed.

We also performed the same analysis using an external cohort, namely the TCGA-SARC cohort. We classified the patients integrated in the TCGA-SARC cohort with SARCULATOR and consequently obtained a 5-year OS probability for each patient. Then, we calculated the C-Indexes (derived from different Cox Proportional Hazards Models for OS) and compared them. We considered different models and distinct model combinations, including SARCULATOR and TC.

CINSARC classification was applied to the TCGA-SARC dataset, based on a previous study that had already classified TCGA-SARC data using CINSARC [50]. Using the available CINSARC code (https://codeocean.com/capsule/4933686/tree/v1), TCGA subtypes (DDLPS, UPS, and LMS) were reclassified according to the CINSARC C1 and C2 categories. Kaplan-Meier curves were then generated to evaluate OS predictions based on CINSARC classification and transcriptomic clusters. Additionally, a Cox Proportional Hazards Model was employed to assess OS, also incorporating other relevant clinical variables for a more comprehensive analysis. Finally, we incorporated CINSARC gene list in the ssGSEA cluster assignment and analyzed the normalized enrichment scores (NES), using Spearman’s rank test to correlate the enrichment scores of each gene set.

As previously described, we performed a comparative analysis, using the TCGA-SARC cohort, of the C-Indexes of different survival estimation models, including SARCULATOR, CINSARC and transcriptomic clusters.

## Data Availability

Different data and generated datasets have been deposited in figshare under the following URL: https://figshare.com/s/6a70cbb12d2738a6e60b (to be made public upon publication).

## Code Availability

All code is available at https://github.com/QuantitativeBiology/Sarcoma-TC-Clusters.

## Data Availability

Data produced in the present work are contained in the manuscript. Extensive data and all generated datasets have been deposited in figshare under the following URL: https://figshare.com/s/6a70cbb12d2738a6e60b (to be made public upon publication).
All code is available at https://github.com/QuantitativeBiology/Sarcoma-TC-Clusters.

https://github.com/QuantitativeBiology/Sarcoma-TC-Clusters.

## Acknowledgements

This research was sponsored by Roche Foundation Medicine under the RNA LDT Research Programme. We would like to thank Roche Foundation Medicine for providing the FoundationOne®CDx and FoundationOne®RNA assays, and for all the technical support, specifically during the transfer of the sequencing data via the safe platform.

We would like to thank Sarah Yacoub (Foundation Medicine, Inc.) for all the precious help in the logistical operationalization of the transfer of all of the different batches of samples between iMM (Lisbon, Lisbon, Portugal) and the Foundation Medicine Headquarters (Cambridge, Massachusetts, United States of America) and for her valuable assistance on sending the results of both DNA and RNAseq results via a safe platform.

We would also like to thank Rachel Beth Keller-Evans (Foundation Medicine, Inc.) for her crucial support in the analysis of the data that has resulted from the employment of the FoundationOne®RNA assay, namely for her help in the analysis of the detected fusions.

We would also like to show our deepest gratitude to the IPOLFG tumor biobank staff, that were responsible for the original retrieval and organization of the FFPE samples that were used in this study, the IPOLFG pathology department staff, and also to the iMM Comparative Pathology unit team (that have sectioned the blocks), the iMM Translational Oncobiology Lab staff (that have helped in the preparation and shipment of the different batches of samples), and the iMM Technology Transfer Office staff.

We would finally like to than Brian Van Tine and Alliny C S Bastos for their exquisite input and feedback.

## Author Information

### Contributions

#### Contribution of each of the participating institutions

This project was developed by Instituto de Medicina Molecular João Lobo Antunes (iMM) - the biomedical research institute whose activity is linked to one of the sarcoma national reference centers in Lisbon, Unidade Local de Saúde Santa Maria (ULSSM), and that is part, together with ULSSM, of the Centro Académico de Medicina de Lisboa (CAML) -, Instituto Português de Oncologia de Lisboa Francisco Gentil (IPOLFG) – a sarcoma international (European) and national reference center in Lisbon - and Instituto Superior Técnico (IST) - the largest school and research institute in Engineering in Portugal - in collaboration with F.Hoffmann-LaRoche and Foundation Medicine, Inc.

The present study was performed in accordance with the ethical standards of Helsinki Declaration II and was approved by the Institution Review Boards of both CAML and IPOLFG.

IPOLFG provided formalin-fixed paraffin-embedded (FFPE) STS samples of 102 patients. iMM prepared and shipped the samples to the Foundation Medicine, Inc-(Cambridge, MA, USA). F.Hoffmann-LaRoche and Foundation Medicine, Inc. provided 100 quantities of FoundationOne®CDx for DNA sequencing (DNAseq) and 100 quantities of FoundationOne®RNA for RNA sequencing (RNAseq), extracted both DNA and RNA from the samples, performed the up mentioned DNAseq and RNAseq and made the subsequent results available for analysis by iMM, IPOLFG and IST.

#### Contribution of each of the authors

MEM, MSR, IF, EG, and HV conceptualized the study. MML selected the FFPE STS sarcoma samples and conducted a detailed anatomopathological review of each one of them. FF, NA and HV retrieved clinical data from the patients whose samples were analyzed and built the clinical database. MEM, DC and SD operationalized the shipment of the samples from iMM to the Foundation Medicine Headquarters. JAL and RH operationalized the sending of the genomic and transcriptomic data resultant from both DNA and RNA sequencing via a safe platform from Foundation Medicine to iMM. MEM, MSR, LGP, EG, and HV implemented analysis, while MSR, LGP and EG ran the unsupervised machine learning-based analysis of the sequencing data. MEM, MSR, IF, JAL, RH, LC, NA, EG and HV contributed to methodology. IF, LC, NA, EG and HV supervised the study. MEM, MSR, FF, EG and HV wrote the manuscript. All authors have revised and approved the manuscript.

#### Ethics Declarations

The present study was performed in accordance with the ethical standards of Helsinki Declaration II and was approved by the Institution Review Boards of both CAML and IPOLFG.

## Notes

### Competing Interest Statement

The authors have declared no competing interest.

### Funding Statement

This study was sponsored by Roche Foundation Medicine under the RNA LDT Research Programme. Roche Foundation Medicine provided all the FoundationOneCDx and FoundationOneRNA assays, and also provided technical support.

### Author Declarations

Ethics committee of Centro Academico de Medicina de Lisboa gave ethical approval for this work. Ethics committee of Instituto Portugues de Oncologia de Lisboa Francisco Gentil gave ethical approval for this work.

