## Supplemental Files for "Machine learning-based analysis of genomic and transcriptomic data unveils sarcoma clusters with superlative prognostic and predictive value"

### Supplementary Material

#### Results

**Genes included in CINSARC 67-gene prognostic signature**

ANLN, *ASPM, AURKA, AURKB, BIRC5, *BORA, BUB1, BUB1B, CCNA2, CCNB1, CCNB2, CDC20, CDC45, CDC6, *CDC7, *CDCA2, CDCA3, CDCA8, *CDK1, CENPA, *CENPE, *CENPL, CEP55, CHEK1, *CKAP5, *CKS2, ECT2, *ESPL1, *FANCI, *FBXO5, FOXM1, H2AFX, *HP1BP3, *KIF11, *KIF14, *KIF15, *KIF18A, *KIF20A, KIF23, KIF2C, *KIF4A, *MAD2L1, MCM2, *MCM7, MELK, *NCAPH, NDE1, NEK2, NUF2, *OIP5, PBK, *PLK4, *PRC1, PTTG1, RAD51AP1, *RNASEH2A, RRM2, *SGO2, *SMC2, *SPAG5, *SPC25, TOP2A, TPX2, TRIP13, TTK, UBE2C, ZWINT

Note: Genes included in the CINSARC 67-gene panel whose expression is not evaluated by F1RNA are marked with an asterisk (*).

**Characterization of frequency and types of genomic alterations detected by FoundationOne®CDx in patients included in each transcriptomic cluster**

Among the patients included in C1, the most frequently found gene alterations were amplifications, especially in the MDM2 gene, whose amplification was found in 15 cases. Amplifications were also frequently found in other genes, such as CDK4 (7 cases) and JUN (6 cases). On the other hand, copy-number-losses in NF1 were found in 4 cases, while short variants in TP53 were identified in 5 cases. The genomic profile of the patients included in C1 is mostly portrayed by a mixture of frequent copy-number-gains in MDM2, CDK4, and JUN, along with copy-number-losses in NF1 and short variants (point mutations) in TP53.

Among the patients that comprise C2, the most commonly identified gene alterations were short variants. TP53 was the gene where alterations were more frequently found (18 cases). ATRX and RB1 alterations were also frequently verified. RB1 copy-number-losses were observed in 10 cases. In parallel, PTEN copy-number losses were verified in 5 cases, and MED12 and NTRK1 alterations were observed in 4 and 3 cases, respectively. The genomic profile of the patients included in C2 is defined by frequent TP53 alterations of different types (mostly short-variants and copy-number-losses) and

RB1 copy-number-losses, with an additional plethora of alterations in ATRX, PTEN, MED12, and NTRK1.

Amidst patients that integrate C3, the pattern of identified genomic alterations was largely marked by deletions (short variants and copy-number-losses), with TP53 displaying a high frequency of short variants (16 cases) (alongside APC (5 cases), MLH1 (4 cases), and RAD54L (3 cases)), while CDKN2A, CDKN2B and MTAP were frequently affected by copy-number losses. Concomitantly, NF1 deletions were observed in 6 cases. The genomic profile of the patients included in C3 is principally portrayed by a high frequency of deletions in CDKN2A/B, MTAP and NF1, alongside frequent point mutations in TP53 and other tumor suppressor genes.

Regarding patients that are part and compose C4, short variants have been often verified, more particularly in TP53 (7 cases) and PTPN11 (3 cases). Copy-number-losses have also been verified in RB1 (6 cases). Finally, FGF23 rearrangements are also common, being observed in 4 cases. The genomic profile of patients included in C4 is characterized by a combination of RB1 copy-number-losses, TP53 mutations (short variants and copy-number-losses), and frequent FGF23 and PTPN11 structural rearrangements.

#### Supplementary Figures

**Supplementary Figure 1 –** Consensus clustering analysis: Optimal number of clusters. Consensus matrix (1a) and Delta area plot (1b).

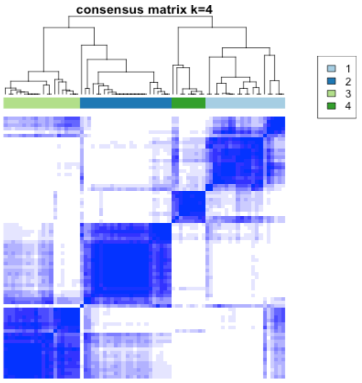

**1a**

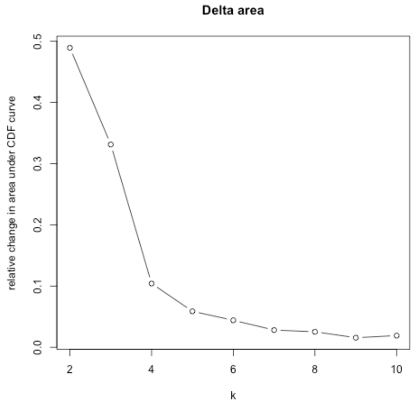

**1b**

**Supplementary Figure 2 -** Comparison of differentially expressed genes between pairs of clusters (Volcano plots)

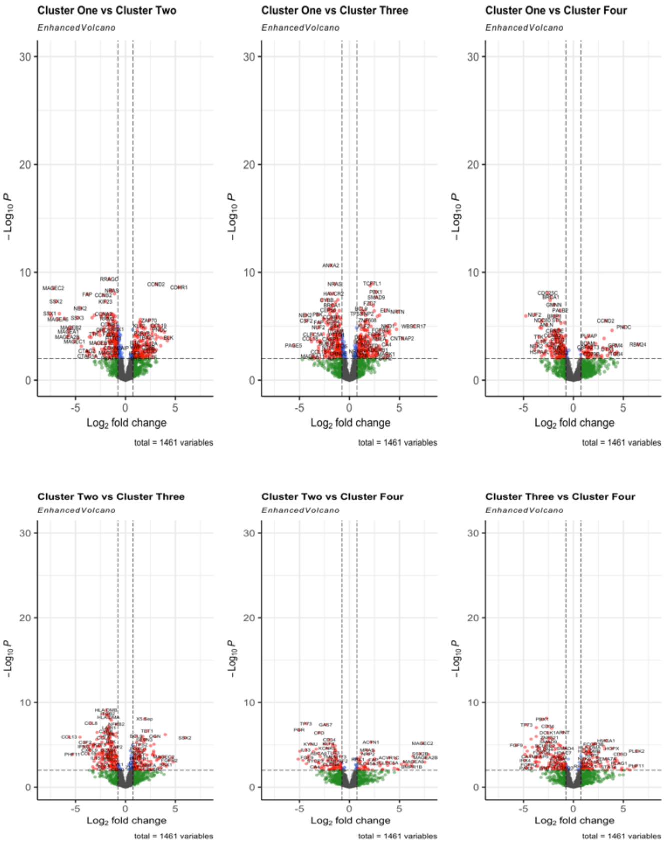

**Supplementary Figure 3 -** ssGSEA Normalized Enrichment Score for each TCGA-SARC patient (Heatmap Plot).

The heatmap plot show a significant enrichment of these patients samples to c1_under and c3_over.

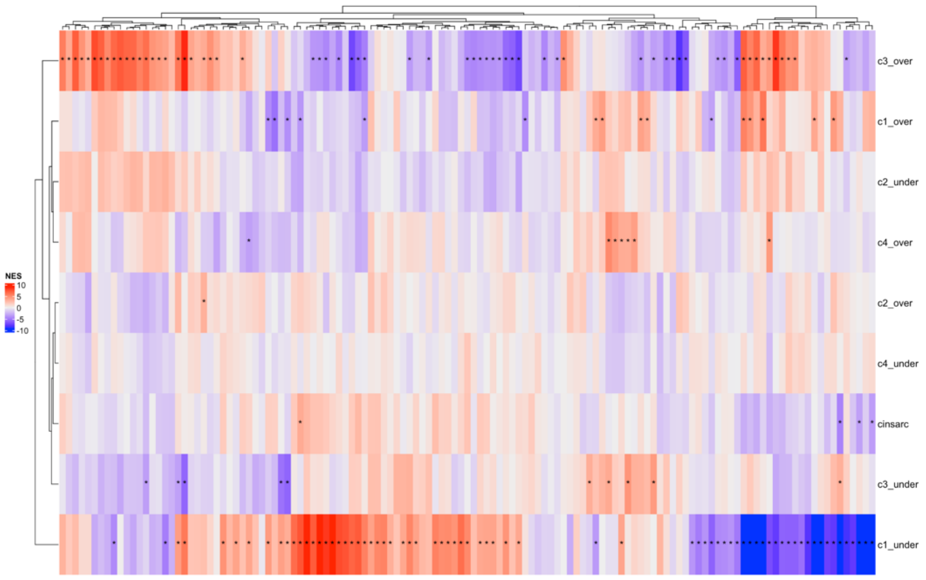

**Supplementary Figure 4 -** Persistence of ssGSEA enrichment in C1_under and C3_over of the TCGA-SARC population persist following the exclusion of UPS patients

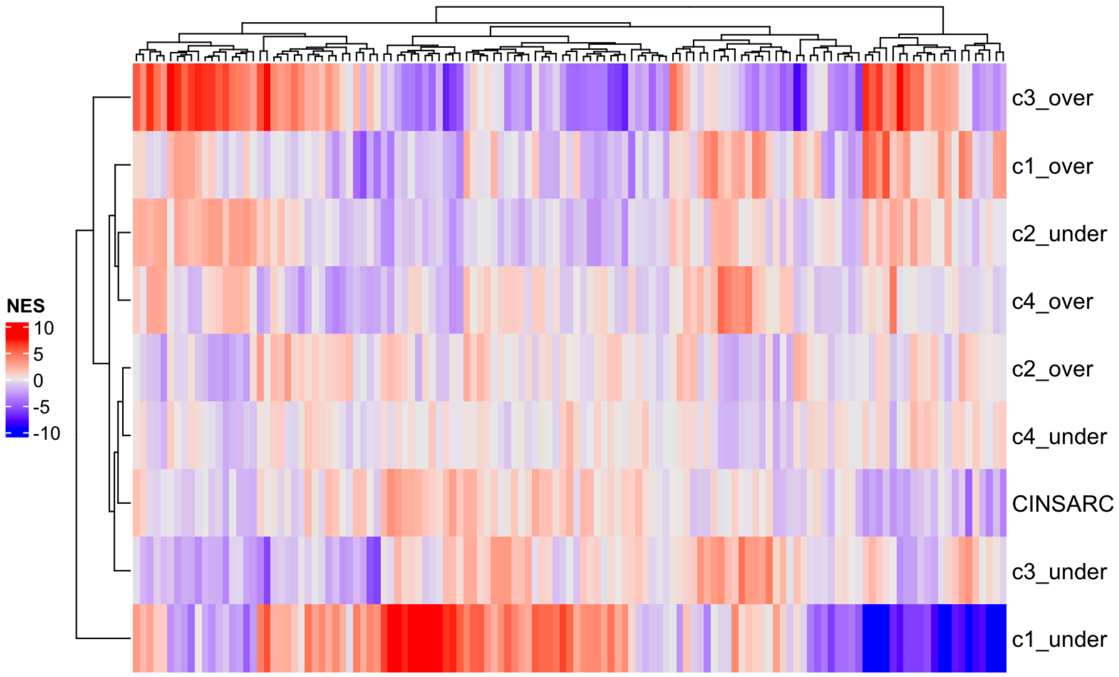

**Supplementary Figure 5** - Differential impact of distinct histopathological and molecular factors on overall survival considering TCGA-SARC patients, after the exclusion of UPS patients (Forest plot).

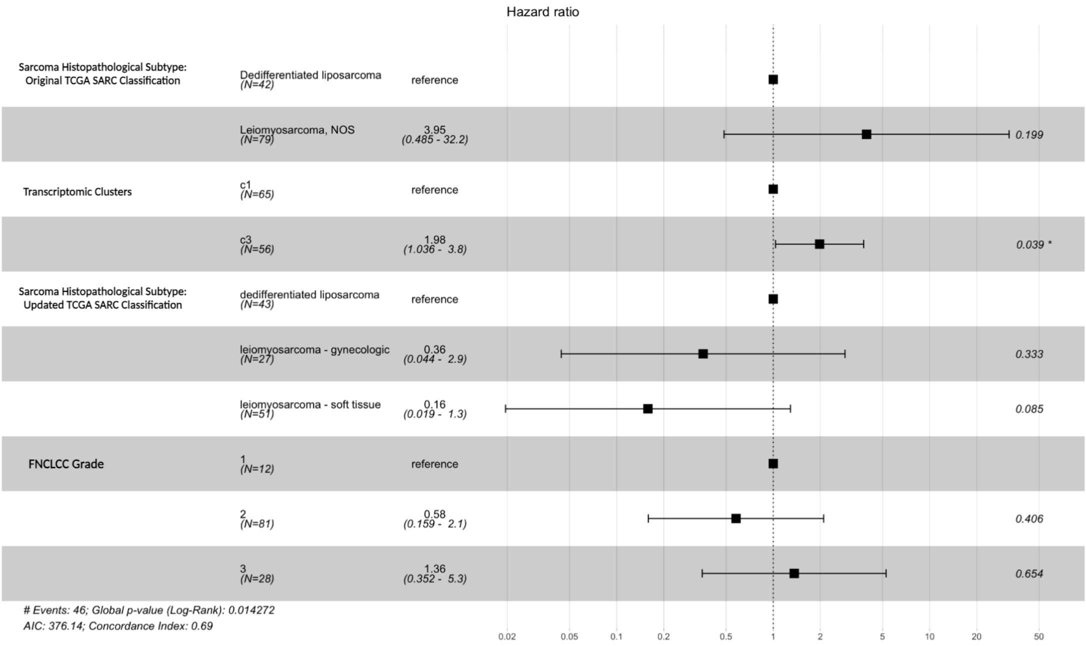

**Supplementary Figure 6 -** Survival analysis (using the Kaplan-Meier method) of the study cohort patients that have displayed either a 5-year OS ≤ 60% and a 5-year OS > 60% (Sarculator-based): OS analysis by the Kaplan-Meier method and respective curves.

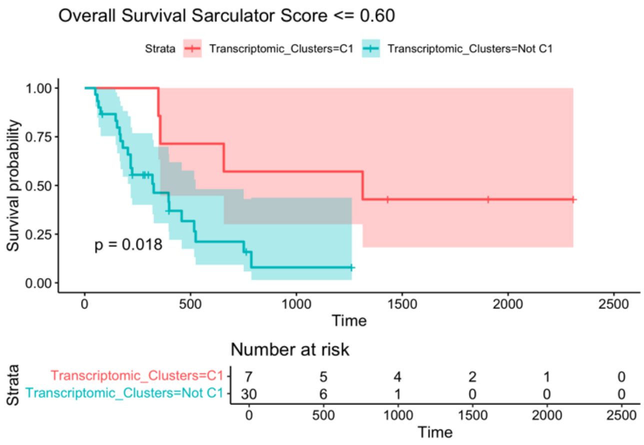

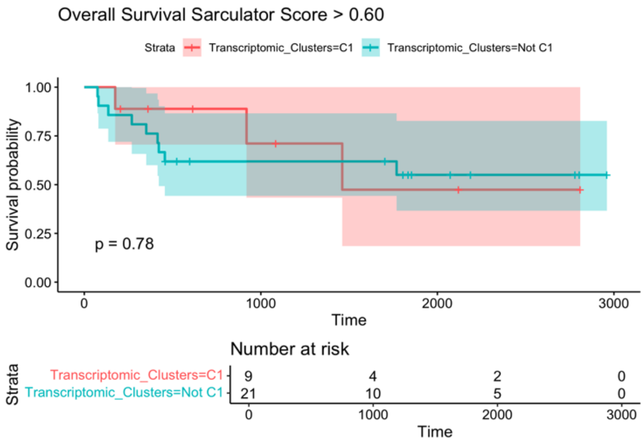

**Supplementary Figure 7 -** Survival analysis (using the Kaplan-Meier method) of the TCGA-SARC (external cohort) patients that have displayed either a 5-year OS ≤ 60% and a 5-year OS > 60% (Sarculator-based): OS analysis by the Kaplan-Meier method and respective curves. The generic analysis including DDLPS, LMS and UPS patients from the TCGA-SARC cohort is displayed on **Supplementary Figure 7a.** The specific analysis including DDLPS, LMS and UPS patients from the TCGA-SARC cohort and that display an STS with an FNLCC grade 3 are shown on **Supplementary Figure 7b.**

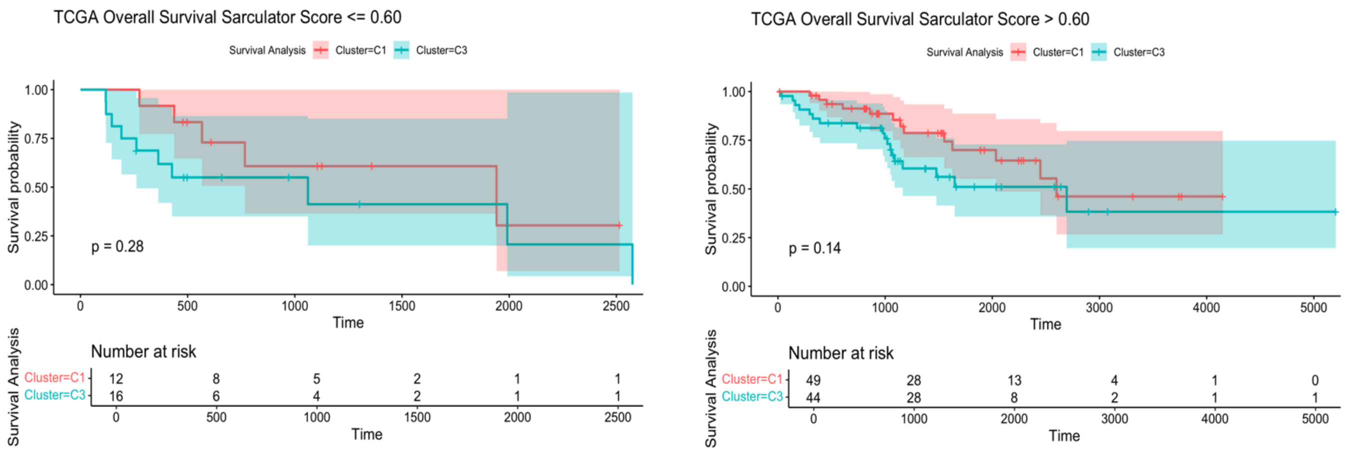

**Supplementary Figure 7a**

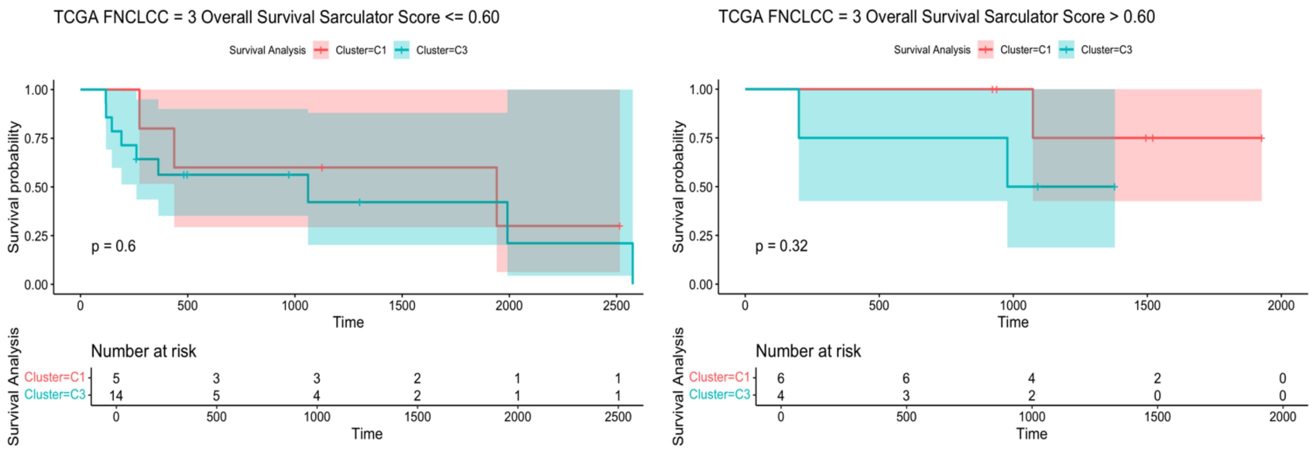

**Supplementary Figure 7b**

**Supplementary Figure 8 -** Correlation between ssGSEA enrichment scores (NES) from c1_under, c3_over and CINSARC genes (Spearman correlation plot).

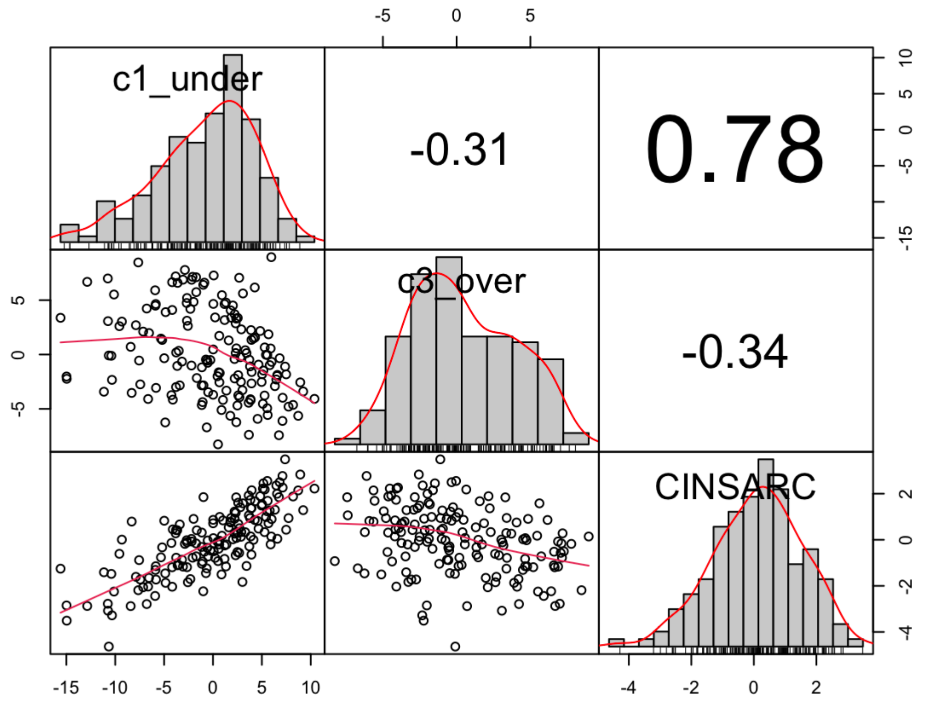

#### Supplementary Tables

**Supplementary Table 1 –** Demographic characteristics of the study population and main features of the included STS cases

| | Total<br>(n=101) |
|---|---|
| **Age at diagnosis**, median [IQR], years | 67 [19.7] |
| **Gender**, n (%) | |
| Male | 50 (49.5) |
| Female | 51 (50.5) |
| **Histopathological subtypes**, n (%) | |
| Dedifferentiated liposarcoma | 25 (24.8) |
| Leiomyosarcoma | 25 (24.8) |
| Undifferentiated pleomorphic sarcoma | 51 (50.4) |
| **Location**, n (%) | |
| Upper limb | 9 (8.9) |
| Lower limb | 49 (48.5) |
| Retroperitoneum | 31 (30.7) |
| Trunk | 12 (11.9) |
| **Presentation type**, n (%) | |
| Localized | 96 (95.0) |
| Distant metastasis | 5 (5.0) |
| **Size of the primary tumor**, median [IQR], cm | 13 [10.0] |
| *Missing* | 4 |

**Supplementary Table 2 –** Surgical and systemic treatment details of the patients who were considered for a curative surgical approach at IPOLFG

| | **Total** (n=94) |
|---|---|
| **Neoadjuvant treatment,** n (%), n=94 | |
| No neoadjuvant treatment | 91 (96.8) |
| Neoadjuvant treatment | 3 (3.2) |
| **Resectability,** n (%), n = 94 | |
| Resectable | 92 (97.9) |
| Unresectable | 2 (2.1) |
| **Resection margins,** n (%), n=92 | |
| R0/R1 | 89 (96.7) |
| R2 | 3 (3.3) |
| **Adjuvant treatment,** n (%), n=92 | |
| Radiotherapy | 55 (59.7) |
| No adjuvant treatment | 34 (37.0) |
| Chemotherapy or Chemoradiotherapy | 3 (3.3) |

**Supplementary Table 3 –** Actionable gene variants (distributed per ESCAT evidence tier) found for patients included in C1.

For each variant, gene identification, nature of the alteration, functional relevance evidence for the alteration (A – Curated; B – Assumed; C - Predicted) and the predictive value of the alteration is provided (2 - Investigational; 3 – Hypothetical target: Alteration-drug match is associated with antitumor activity, but magnitude of benefit is unknown (potential cancer-repurposing opportunity); 4 – Hypothetical target: pre-clinical evidence of actionability).

| Tier | Gene | Alteration | Functional relevance evidence | Biomarker predictive value |
|---|---|---|---|---|
| **2** | MDM 2 | Copy number alteration (Amplification) | Evidence A Oncogenic | Sensitivity/Response:<br>- Brigimadlin.<br>- Milademetan. |
| | MTAP | Copy number alteration (Deletion) | Evidence A Likely oncogenic | Sensitivity/Response:<br>- MRTX1719.<br>- AMG193. |
| | TP53 | Mutation – missense p.Tyr234Asp exon 7/11 | Evidence A Pathogenic / Likely Pathogenic | Sensitivity/Response:<br>- Pazopanib.<br>- Vorinostat. |
| | TP53 | Mutation – frameshift p.Glu339Ter exon 10/11 | Evidence A Pathogenic | Sensitivity/Response:<br>- Pazopanib.<br>- Vorinostat. |
| | TP53 | Mutation - stop gained p.Arg196Ter exon 6/11 | Evidence A Pathogenic | Sensitivity/Response:<br>- Pazopanib.<br>- Vorinostat. |
| | TP53 | Mutation – missense p.Met246Lys exon 7/11 | Evidence A Likely Pathogenic | Sensitivity/Response:<br>- Pazopanib.<br>- Vorinostat. |
| **3** | MDM2 | Copy number alteration (Amplification) | Evidence A Oncogenic | Sensitivity/Response:<br>- Brigimadlin.<br>- Ezabenlimab. |
| | NF1 | Mutation - stop gained p.Gln1870Ter exon 38/57 | Evidence A Pathogenic | Sensitivity/Response:<br>- Selumetinib.<br>Resistance/Reduced Sensitivity:<br>- Vemurafenib. |
| | TP53 | Mutation – missense p.Tyr234Asp exon 7/11 | Evidence A Pathogenic / Likely Pathogenic Likely Oncogenic | Sensitivity/Response:<br>- Azacitidne.<br>- Eprenetapopt.<br>- Chemotherapy.<br>Resistance/Reduced Sensitivity:<br>- RG7112. |
| | TP53 | Mutation – frameshift | Evidence A Pathogenic | Sensitivity/Response:<br>- Azacitidne. |

| | | | | |
|---|---|---|---|---|
| | | p.Glu339Ter exon 10/11 | | - Eprenetapopt.<br>- Chemotherapy.<br>Resistance/Reduced Sensitivity:<br>- RG7112. |
| | TP53 | Mutation - stop gained p.Arg196Ter exon 6/11 | Evidence A Pathogenic | Sensitivity/Response:<br>- Azacitidne.<br>- Eprenetapopt.<br>- Chemotherapy.<br>Resistance/Reduced Sensitivity:<br>- RG7112. |
| | TP53 | Mutation – missense p.Met246Lys exon 7/11 | Evidence A Likely Pathogenic Likely Oncogenic | Sensitivity/Response:<br>- Azacitidne.<br>- Eprenetapopt.<br>- Chemotherapy.<br>Resistance/Reduced Sensitivity:<br>- RG7112. |
| | RAD51B | Mutation – frameshift p.Leu31PhefsTer9 exon 3/11 | Evidence B | Sensitivity/Response:<br>- Olaparib. |
| | NF1 | Mutation – frameshift p.Gln2702HisfsTer6 exon 56/57 | Evidence B | Sensitivity/Response:<br>- Selumetinib.<br>Resistance/Reduced Sensitivity:<br>- Vemurafenib. |
| | ATM | Mutation – missense p.Ser2408Leu exon 49/63 | Evidence C | Sensitivity/Response:<br>- Olaparib.<br>- Talazoparib. |
| | BRIP1 | Mutation – missense p.Arg264Trp exon 7/20 | Evidence C | Sensitivity/Response:<br>- Olaparib. |
| **4** | CDK4 | Copy number alteration (Amplification) | Evidence A Oncogenic | Sensitivity/Response:<br>- Palbociclib. |
| | MDM2 | Copy number alteration (Amplification) | Evidence A Oncogenic | Sensitivity/Response:<br>- Brigimadlin.<br>- Milademetan. |
| | TP53 | Mutation - stop gained p.Arg196Ter exon 6/11 | Evidence A Pathogenic | Resistance/Reduced Sensitivity:<br>- Rebemadlin.<br>- MDM2 inhibitor AMGMDS3. |
| | TP53 | Mutation – missense p.Met246Lys exon 7/11 | Evidence A Likely pathogenic Likely oncogenic | Resistance/Reduced Sensitivity:<br>- Rebemadlin.<br>- MDM2 inhibitor AMGMDS3. |
| | TP53 | Mutation – missense p.Tyr234Asp exon 7/11 | Evidence A Likely pathogenic Likely oncogenic | Resistance/Reduced Sensitivity:<br>- Rebemadlin.<br>- MDM2 inhibitor AMGMDS3. |

| | | | | |
|---|---|---|---|---|
| | TP53 | Mutation – frameshift p.Glu339Ter exon 10/11 | Evidence A Pathogenic | Resistance/Reduced Sensitivity:<br>- Rebemadlin.<br>- MDM2 inhibitor AMGMDS3. |
| | NF1 | Mutation - stop gained p.Gln1870Ter exon 38/57. | Evidence A Pathogenic | Sensitivity/Response:<br>- Trametinib.<br>- Cobimetinib. |
| | NF1 | Mutation – frameshift p.Gln2702HisfsTer6 exon 56/57 | Evidence B | Sensitivity/Response:<br>- Trametinib.<br>- Cobimetinib |

Note: It is important to underline an important number of alterations without a specific predictive value, but with clear functional relevance evidence of the genes AMER1, NTRK1, PTEN, PI3KCA, RB1.

**Supplementary Table 4 -** Actionable gene variants (distributed per ESCAT evidence tier) found for patients included in C2.

For each variant, gene identification, nature of the alteration, functional relevance evidence for the alteration (A – Curated; B – Assumed; C - Predicted) and the predictive value of the alteration is provided (2 - Investigational; 3 – Hypothetical target: Alteration-drug match is associated with antitumor activity, but magnitude of benefit is unknown (potential cancer-repurposing opportunity); 4 – Hypothetical target: pre-clinical evidence of actionability).

| Tier | Gene | Alteration | Functional relevance evidence | Biomarker predictive value |
|---|---|---|---|---|
| **2** | MDM 2 | Copy number alteration (Amplification) | Evidence A<br>Oncogenic | Sensitivity/Response:<br>- Brigimadlin.<br>- Milademetan. |
| | TP53 | Mutation – missense p.Val157Phe exon 5/11 | Evidence A<br>Likely oncogenic<br>Resistance | Sensitivity/Response:<br>- Pazopanib.<br>- Vorinostat. |
| | TP53 | Mutation - stop gained p.Gln144Ter exon 5/11 | Evidence A<br>Pathogenic | Sensitivity/Response:<br>- Pazopanib.<br>- Vorinostat. |
| | TP53 | Mutation – missense p.Ile195Thr exon 6/11 | Evidence A<br>Likely oncogenic | Sensitivity/Response:<br>- Pazopanib.<br>- Vorinostat. |
| | TP53 | Mutation – frameshift p.Arg209LysfsTer6 exon 6/11 | Evidence A<br>Pathogenic | Sensitivity/Response:<br>- Pazopanib.<br>- Vorinostat. |
| | TP53 | Mutation – missense p.Tyr126Asn exon 5/11 | Evidence A<br>Likely oncogenic | Sensitivity/Response:<br>- Pazopanib.<br>- Vorinostat. |
| | TP53 | Mutation - stop gained p.Arg306Ter exon 8/11 | Evidence A<br>Pathogenic | Sensitivity/Response:<br>- Pazopanib.<br>- Vorinostat. |
| | TP53 | Mutation - stop gained p.Gln144Ter exon 5/11 | Evidence A<br>Pathogenic | Sensitivity/Response:<br>- Pazopanib.<br>- Vorinostat. |
| | MTAP | Copy number alteration (Deletion) | Evidence A<br>Likely oncogenic | Sensitivity/Response:<br>- MRTX1719.<br>- AMMG193. |
| | ERBB2 | Copy number alteration (Amplification) | Evidence A<br>Oncogenic | Sensitivity/Response:<br>- Trastuzumab Deruxtecan. |
| | TP53 | Mutation – frameshift p.His178ProfsTer69 exon 5/11 | Evidence B | Sensitivity/Response:<br>- Pazopanib.<br>- Vorinostat. |

| | | | | |
|---|---|---|---|---|
| | TP53 | Mutation - stop gained p.Cys182Ter exon 5/11 | Evidence B | Sensitivity/Response:<br>- Pazopanib.<br>- Vorinostat. |
| | TSC2 | Mutation – frameshift p.Gly654ValfsTer2 exon 19/42 | Evidence B | Sensitivity/Response:<br>- ABI-009. |
| | TSC2 | Mutation – missense p.Arg59Gln exon 3/42 | Evidence C | Sensitivity/Response:<br>- ABI-009. |
| **3** | FGFR1 | Mutation – missense p.Asn577Lys exon 13/19 | Evidence A<br>Pathogenic<br>Likely oncogenic<br>Sensitivity/Response | Sensitivity/Response:<br>- Pemigatinib. |
| | MDM2 | Copy number alteration (amplification) | Evidence A<br>Oncogenic | Sensitivity/Response:<br>- Brigimadlin.<br>- Ezabenlimab. |
| | RET | Mutation – missense p.Val648Ile exon 11/20 | Evidence A<br>Likely oncogenic | Sensitivity/Response:<br>- Selpercatinib.<br>- Pralsetinib. |
| | ERBB2 | Copy number alteration (amplification) | Evidence A<br>Oncogenic | Sensitivity/Response:<br>- Trastuzumab.<br>- Trastuzumab, Pertuzumab.<br>- Trastuzumab, Chemotherapy.<br>- Ado-Trastuzumab Emtansine.<br>- Trastuzumab Deruxtecan.<br>- Tucatinib, Trastuzumab.<br>- Tucatinib, Trastuzumab, Capecitabine.<br>- Lapatinib, Capecitabine.<br>- Lapatinib, Trastuzumab.<br>- Neratinib.<br>- Neratinib, Capecitabine.<br>- Margetuximab, Chemotherapy. |
| | NF1 | Mutation - stop gained p.Trp571Ter exon 15/57 | Evidence A<br>Pathogenic | Sensitivity/Response:<br>- Selumetinib.<br>Resistance/Reduced Sensitivity:<br>- Vemurafenib. |
| | POLE | Mutation – missense p.Gly6Arg exon 1/49 | Evidence A<br>Likely oncogenic | Sensitivity/Response:<br>- Pembrolizumab. |

| | | | | |
|---|---|---|---|---|
| | TP53 | Mutation – missense p.Val157Phe exon 5/11 | Evidence A<br>Likely oncogenic<br>Resistance | Sensitivity/Response:<br>- Azacitidne.<br>- Eprenetapopt.<br>- Chemotherapy.<br>Resistance/Reduced Sensitivity:<br>- RG7112. |
| | TP53 | Mutation - stop gained p.Gln144Ter exon 5/11 | Evidence A<br>Pathogenic | Sensitivity/Response:<br>- Azacitidne.<br>- Eprenetapopt.<br>- Chemotherapy.<br>Resistance/Reduced Sensitivity:<br>- RG7112. |
| | TP53 | Mutation – missense p.Ile195Thr exon 6/11 | Evidence A<br>Likely oncogenic | Sensitivity/Response:<br>- Azacitidne.<br>- Eprenetapopt.<br>- Chemotherapy.<br>Resistance/Reduced Sensitivity:<br>- RG7112. |
| | TP53 | Mutation – missense p.Tyr126Asn exon 5/11 | Evidence A<br>Likely oncogenic | Sensitivity/Response:<br>- Azacitidne.<br>- Eprenetapopt.<br>- Chemotherapy.<br>Resistance/Reduced Sensitivity:<br>- RG7112. |
| | TP53 | Mutation - stop gained p.Arg306Ter exon 8/11 | Evidence A<br>Pathogenic | Sensitivity/Response:<br>- Azacitidne.<br>- Eprenetapopt.<br>- Chemotherapy.<br>Resistance/Reduced Sensitivity:<br>- RG7112. |
| | TP53 | Mutation - stop gained p.Gln144Ter exon 5/11 | Evidence A<br>Pathogenic | Sensitivity/Response:<br>- Azacitidne.<br>- Eprenetapopt.<br>- Chemotherapy.<br>Resistance/Reduced Sensitivity:<br>- RG7112. |
| | PTEN | Mutation – frameshift p.Ile101LysfsTer5 exon 5/9 | Evidence B | Sensitivity/Response:<br>- Fulvestrant.<br>- Capivasertib. |
| | PTEN | Mutation – frameshift p.Glu40ValfsTer20 exon 2/9 | Evidence B | Sensitivity/Response:<br>- Fulvestrant.<br>- Capivasertib. |
| **4** | TP53 | Mutation – frameshift p.His178ProfsTer69 exon 5/11 | Evidence B | Sensitivity/Response:<br>- Azacitidne.<br>- Eprenetapopt.<br>- Chemotherapy.<br>Resistance/Reduced Sensitivity:<br>- RG7112. |

| | | | | |
|---|---|---|---|---|
| | TP53 | Mutation - stop gained p.Cys182Ter exon 5/11 | Evidence B | Sensitivity/Response:<br>- Azacitidne.<br>- Eprenetapopt.<br>- Chemotherapy.<br>Resistance/Reduced Sensitivity:<br>- RG7112. |
| | RAD51C | Mutation – frameshift p.Val298AlafsTer6 exon 6/9 | Evidence B | Sensitivity/Response:<br>- Olaparib.<br>- Talazoparib. |
| | TSC2 | Mutation – frameshift p.Gly654ValfsTer2 exon 19/42 | Evidence B | Sensitivity/Response:<br>- Everolimus.<br>- ABI-009. |
| | TSC2 | Mutation – missense p.Arg59Gln exon 3/42 | Evidence C | Sensitivity/Response:<br>- Everolimus.<br>- ABI-009. |
| | CDK4 | Copy number alteration (Amplification) | Evidence A<br>Oncogenic | Sensitivity/Response:<br>- Palbociclib.<br>- Abemaciclib. |
| | FGFR1 | Mutation – missense p.Asn577Lys exon 13/19 | Evidence A<br>Pathogenic<br>Likely oncogenic<br>Sensitivity/Response | Sensitivity/Response:<br>- AZD4547.<br>- Erdafitinib. |
| | MDM2 | Copy number alteration (Amplificaton) | Evidence A<br>Oncogenic. | Sensitivity/Response:<br>- Brigimadlin.<br>- Milademetan. |
| | TP53 | Mutation – missense p.Val157Phe exon 5/11 | Evidence A<br>Likely oncogenic.<br>Resistance. | Resistance/Reduced Sensitivity:<br>- Rebemadlin.<br>- MDM2 inhibitor AMGMDS3. |
| | TP53 | Mutation - stop gained p.Gln144Ter exon 5/11 | Evidence A<br>Pathogenic. | Resistance/Reduced Sensitivity:<br>- Rebemadlin.<br>- MDM2 inhibitor AMGMDS3. |
| | TP53 | Mutation – missense p.Ile195Thr exon 6/11 | Evidence A<br>Likely oncogenic | Resistance/Reduced Sensitivity:<br>- Rebemadlin.<br>- MDM2 inhibitor AMGMDS3. |
| | TP53 | Mutation – frameshift p.Arg209LysfsTer6 exon 6/11 | Evidence A<br>Pathogenic | Resistance/Reduced Sensitivity:<br>- Rebemadlin.<br>- MDM2 inhibitor AMGMDS3. |
| | TP53 | Mutation – missense p.Tyr126Asn exon 5/11 | Evidence A<br>Likely oncogenic. | Resistance/Reduced Sensitivity:<br>- Rebemadlin.<br>- MDM2 inhibitor AMGMDS3. |
| | TP53 | Mutation - stop gained p.Arg306Ter exon 8/11 | Evidence A<br>Pathogenic. | Resistance/Reduced Sensitivity:<br>- Rebemadlin. |

| | | | | |
|---|---|---|---|---|
| | | | | - MDM2 inhibitor AMGMDS3. |
| | TP53 | Mutation - stop gained p.Gln144Ter exon 5/11 | Evidence A Pathogenic | Resistance/Reduced Sensitivity:<br>- Rebemadlin.<br>- MDM2 inhibitor AMGMDS3. |
| | CCNE1 | Copy number alteration (Amplification) | Evidence A Oncogenic. | Sensitivity/Response:<br>- RP-6306. |
| | NF1 | Mutation - stop gained p.Trp571Ter exon 15/5 | Evidence A Pathogenic. | Sensitivity/Response:<br>- Trametinib.<br>- Cobimetinib. |
| | PTEN | Mutation – frameshift p.Glu40ValfsTer20 exon 2/9 | Evidence B | Sensitivity/Response:<br>- GSK2636771.<br>- AZD8186.<br>- Ipatasertib. |
| | PTEN | Mutation – frameshift p.Ile101LysfsTer5 exon 5/9 | Evidence B | Sensitivity/Response:<br>- GSK2636771.<br>- AZD8186.<br>- Ipatasertib. |
| | TP53 | Mutation – frameshift p.His178ProfsTer69 exon 5/11 | Evidence B | Resistance/Reduced Sensitivity:<br>- Rebemadlin.<br>- MDM2 inhibitor AMGMDS3. |
| | TP53 | Mutation - stop gained p.Cys182Ter exon 5/11 | Evidence B | Resistance/Reduced Sensitivity:<br>- Rebemadlin.<br>- MDM2 inhibitor AMGMDS3. |

Note: It is important to underline an important number of alterations without a specific predictive value, but with clear functional relevance evidence of the genes IGF1R, MUTYH, AURKA, AURKB, BRD4, NTRK1, VEGF, HGF.

**Supplementary Table 5 -** Actionable gene variants (distributed per ESCAT evidence tier) found for patients included in C3.

For each variant, gene identification, nature of the alteration, functional relevance evidence for the alteration (A – Curated; B – Assumed; C - Predicted) and the predictive value of the alteration is provided (2 - Investigational; 3 – Hypothetical target: Alteration-drug match is associated with antitumor activity, but magnitude of benefit is unknown (potential cancer-repurposing opportunity); 4 – Hypothetical target: pre-clinical evidence of actionability).

| Tier | Gene | Alteration | Functional relevance evidence | Biomarker predictive value |
|---|---|---|---|---|
| **2** | MDM 2 | Copy number alteration (Amplification) | Evidence A Oncogenic | Sensitivity/Response:<br>- Brigimadlin.<br>- Milademetan. |
| | MTAP | Copy number alteration (Deletion) | Evidence A Likely oncogenic | Sensitivity/Response:<br>- MRTX1719.<br>- AMG193. |
| | TP53 | Mutation – missense p.His193Tyr exon 6/11 | Evidence A Pathogenic / Likely Pathogenic Likely oncogenic | Sensitivity/Response:<br>- Pazopanib.<br>- Vorinostat. |
| | TP53 | Mutation – missense p.Ala159Val exon 5/11 | Evidence A Pathogenic Likely pathogenic Likely oncogenic | Sensitivity/Response:<br>- Pazopanib.<br>- Vorinostat. |
| | TP53 | Mutation – missense p.Val173Leu exon 5/11 | Evidence A Pathogenic Likely pathogenic Likely oncogenic | Sensitivity/Response:<br>- Pazopanib.<br>- Vorinostat. |
| | TP53 | Mutation – missense p.Arg175His exon 5/1 | Evidence A Pathogenic Oncogenic Poor outcome | Sensitivity/Response:<br>- Pazopanib.<br>- Vorinostat. |
| | TP53 | Mutation – missense p.Arg181Cys exon 5/1 | Evidence A Likely oncogenic | Sensitivity/Response:<br>- Pazopanib.<br>- Vorinostat. |
| | TP53 | Mutation – missense p.Arg248Trp exon 7/1 | Evidence A Pathogenic Likely oncogenic | Sensitivity/Response:<br>- Pazopanib.<br>- Vorinostat. |
| | TP53 | Mutation – missense | Evidence A Pathogenic | Sensitivity/Response:<br>- Pazopanib.<br>- Vorinostat. |

| | | | | |
|---|---|---|---|---|
| | | p.Arg273Cys exon 8/11 | Likely pathogenic | |
| | TP53 | Mutation – missense p.Arg282Gly exon 8/11 | Evidence A Pathogenic Likely oncogenic | Sensitivity/Response:<br>- Pazopanib.<br>- Vorinostat. |
| | TP53 | Mutation – missense p.Glu286Lys exon 8/11 | Evidence A Pathogenic Likely pathogenic Likely oncogenic | Sensitivity/Response:<br>- Pazopanib.<br>- Vorinostat. |
| | TSC2 | Mutation – missense p.Arg1713His exon 40/42 | Evidence A Pathogenic Likely pathogenic | Sensitivity/Response:<br>- ABI-009. |
| | PIK3CA | Mutation – missense p.Ala1066Val exon 21/21 | Evidence A Likely oncogenic | Sensitivity/Response:<br>- Capivasertib.<br>- Copanlisib. |
| **3** | MDM2 | Copy number alteration (Amplification) | Evidence A Oncogenic | Sensitivity/Response:<br>- Brigimadlin.<br>- Ezabenlimab. |
| | POLE | Mutation – missense p.Gly6Arg exon 1/49 | Evidence A Likely oncogenic | Sensitivity/Response:<br>- Pembrolizumab. |
| | POLE | Mutation – missense p.Gly6Arg exon 1/49 | Evidence A Likely oncogenic | Sensitivity/Response:<br>- Pembrolizumab. |
| | TSC2 | Mutation – missense p.Arg1713His exon 40/42 | Evidence A Pathogenic Likely pathogenic | Sensitivity/Response:<br>- Everolimus.<br>- ABI-009. |
| | ATM | Mutation – missense p.Arg337His exon 8/63 | Evidence A Likely oncogenic | Sensitivity/Response:<br>- Olaparib.<br>- Talazoparib. |
| | KRAS | Mutation – missense p.Thr20Ala exon 2/5 | Evidence A Resistance Poor outcome | Sensitivity/Response:<br>- Cobimetinib.<br>- Trametinib.<br>- Binimetinib.<br>- Pemetrexed, Trametinib, Docetaxel.<br>- Docetaxel, Selumetinib.<br>- Abemaciclib.<br>- Nivolumab, Atezolizumab.<br>Resistance/Reduced Sensitivity:<br>- Panitumumab.<br>- Tucatinib, Trastuzumab. |

| | | | | |
|---|---|---|---|---|
| | | | | - Cetuximab.<br>- Panitumumab.<br>- Gemcitabine, Erlotinib.<br>- Gefitinib.<br>- Bevacizumab. |
| | KRAS | Mutation – missense p.Leu19Phe exon 2/5 | Evidence A<br>Oncogenic<br>Resistance<br>Poor outcome | Sensitivity/Response:<br>- Cobimetinib.<br>- Trametinib.<br>- Binimetinib.<br>- Docetaxel, Selumetinib.<br>- Pemetrexed, Trametinib, Docetaxel.<br>- Abemaciclib.<br>- Nivolumab, Atezolizumab.<br>Resistance/Reduced Sensitivity:<br>- Panitumumab.<br>- Tucatinib, Trastuzumab.<br>- Cetuximab.<br>- Panitumumab.<br>- Gemcitabine, Erlotinib.<br>- Gefitinib.<br>- Bevacizumab. |
| | NRAS | Mutation – missense p.Gln61Lys exon 3/7 | Evidence A<br>Oncogenic<br>Resistance | Sensitivity/Response:<br>- Cobimetinib.<br>- Trametinib.<br>- Binimetinib.<br>- Selumetinib.<br>Resistance/Reduced Sensitivity:<br>- Panitumumab.<br>- Tucatinib, Trastuzumab.<br>- Cetuximab.<br>- Panitumumab.<br>- Vemurafenib. |
| | MET | Copy number alteration (Amplification) | Evidence A<br>Oncogenic | Sensitivity/Response:<br>- Telisotuzumab Vedotin.<br>- Tepotinib.<br>- Capmatinib.<br>- Crizotinib.<br>Resistance/Reduced Sensitivity:<br>- Osimertinib.<br>- Gefitinib.<br>- Erlotinib. |
| | PIK3CA | Mutation - missense p.Ala1066Val exon 21/21 | Evidence A<br>Likely oncogenic | Sensitivity/Response:<br>- Capivasertib, Fulvestrant.<br>- Alpelisib, Fulvestrant. |

| | | | | |
|---|---|---|---|---|
| | | | | - Everolimus.<br>Resistance/Reduced Sensitivity:<br>- Trastuzumab.<br>- Lapatinib, Capecitabine.<br>- Cetuximab.<br>- Panitumumab.<br>- Erlotinib.<br>- Gefitinib. |
| | PTEN | Mutation – frameshift p.Lys267ArgfsTer9 exon 7/9 | Evidence A Pathogenic. | Sensitivity/Response:<br>- Capivasertib, Fulvestrant. |
| | TP53 | Mutation – missense p.His193Tyr exon 6/11 | Evidence A Pathogenic Likely pathogenic Likely oncogenic | Sensitivity/Response:<br>- Azacytidine.<br>- Eprenetapopt.<br>- Chemotherapy.<br>Resistance/Reduced Sensitivity:<br>- RG7112. |
| | TP53 | Mutation – missense p.Val173Leu exon 5/11 | Evidence A Pathogenic Likely pathogenic Likely oncogenic | Sensitivity/Response:<br>- Azacytidine.<br>- Eprenetapopt.<br>- Chemotherapy.<br>Resistance/Reduced Sensitivity:<br>- RG7112. |
| | TP53 | Mutation – missense p.Arg175His exon 5/11 | Evidence A Pathogenic Oncogenic | Sensitivity/Response:<br>- Azacytidine.<br>- Eprenetapopt.<br>- Chemotherapy.<br>Resistance/Reduced Sensitivity:<br>- RG7112. |
| | TP53 | Mutation – missense p.Arg181Cys exon 5/11 | Evidence A Likely oncogenic | Sensitivity/Response:<br>- Azacytidine.<br>- Eprenetapopt.<br>- Chemotherapy.<br>Resistance/Reduced Sensitivity:<br>- RG7112. |
| | TP53 | Mutation – missense p.Arg248Trp exon 7/11 | Evidence A Pathogenic Likely oncogenic | Sensitivity/Response:<br>- Azacytidine.<br>- Eprenetapopt.<br>- Chemotherapy.<br>Resistance/Reduced Sensitivity:<br>- RG7112. |
| | TP53 | Mutation – missense p.Arg273Cys exon 8/11 | Evidence A Pathogenic Likely pathogenic Likely oncogenic | Sensitivity/Response:<br>- Azacytidine.<br>- Eprenetapopt.<br>- Chemotherapy.<br>Resistance/Reduced Sensitivity:<br>- RG7112. |

| | | | | |
|---|---|---|---|---|
| | TP53 | Mutation – missense p.Arg282Gly exon 8/11 | Evidence A Pathogenic Likely oncogenic | Sensitivity/Response:<br>- Azacytidine.<br>- Eprenetapopt.<br>- Chemotherapy.<br>Resistance/Reduced Sensitivity:<br>- RG7112. |
| | TP53 | Mutation – missense p.Glu286Lys exon 8/11 | Evidence A Pathogenic Likely pathogenic Likely oncogenic | Sensitivity/Response:<br>- Azacytidine.<br>- Eprenetapopt.<br>- Chemotherapy.<br>Resistance/Reduced Sensitivity:<br>- RG7112. |
| | NF1 | Mutation – frameshift p.Tyr930PhefsTer8 exon 21/57 | Evidence B | Sensitivity/Response:<br>- Selumetinib.<br>Resistance/Reduced Sensitivity:<br>- Vemurafenib. |
| | ATM | Mutation – missense p.Arg2854Cys exon 58/63 | Evidence C | Sensitivity/Response:<br>- Olaparib.<br>- Talazoparib. |
| | ATR | Mutation – missense p.Ser1154Thr exon 18/47 | Evidence C | Sensitivity/Response:<br>- Talazoparib. |
| | VHL | Mutation – missense p.Lys196Glu exon 3/3 | Evidence C | Sensitivity/Response:<br>- Everolimus. |
| **4** | CDK4 | Copy number alteration (Amplification) | Evidence A Oncogenic | Sensitivity/Response:<br>- Abemaciclib.<br>- Palbociclib. |
| | MDM2 | Copy number alteration (Amplification) | Evidence A Oncogenic | Sensitivity/Response:<br>- Brigimadlin.<br>- Milademetan. |
| | CDKN2A | Mutation - stop gained p.Arg58Ter exon 2/3 | Evidence A Pathogenic | Sensitivity/Response:<br>- Abemaciclib.<br>- Palbociclib.<br>- Ribociclib. |
| | TP53 | Mutation – missense p.His193Tyr exon 6/11 | Evidence A Pathogenic Likely pathogenic Likely oncogenic | Resistance/Reduced Sensitivity:<br>- Rebemadlin.<br>- MDM2 inhibitor AMGMDS2. |
| | TP53 | Mutation – missense p.Ala159Val exon 5/11 | Evidence A Pathogenic Likely pathogenic Likely oncogenic | Resistance/Reduced Sensitivity:<br>- Rebemadlin.<br>- MDM2 inhibitor AMGMDS2. |
| | TP53 | Mutation – missense p.Val173Leu exon 5/11 | Evidence A Pathogenic Likely pathogenic | Resistance/Reduced Sensitivity:<br>- Rebemadlin. |

| | | | | |
|---|---|---|---|---|
| | | | Likely oncogenic | - MDM2 inhibitor AMGMDS2. |
| | TP53 | Mutation – missense p.Arg175His exon 5/11 | Evidence A<br>Pathogenic<br>Oncogenic | Resistance/Reduced Sensitivity:<br>- Rebemadlin.<br>- MDM2 inhibitor AMGMDS2. |
| | TP53 | Mutation – missense p.Arg181Cys exon 5/11 | Evidence A<br>Likely oncogenic | Resistance/Reduced Sensitivity:<br>- Rebemadlin.<br>- MDM2 inhibitor AMGMDS2. |
| | TP53 | Mutation – missense p.Arg248Trp exon 7/11 | Evidence A<br>Pathogenic<br>Likely oncogenic | Resistance/Reduced Sensitivity:<br>- Rebemadlin.<br>- MDM2 inhibitor AMGMDS2. |
| | TP53 | Mutation – missense p.Arg273Cys exon 8/11 | Evidence A<br>Pathogenic<br>Likely pathogenic<br>Likely oncogenic | Resistance/Reduced Sensitivity:<br>- Rebemadlin.<br>- MDM2 inhibitor AMGMDS2. |
| | TP53 | Mutation – missense p.Arg282Gly exon 8/11 | Evidence A<br>Pathogenic<br>Likely oncogenic | Resistance/Reduced Sensitivity:<br>- Rebemadlin.<br>- MDM2 inhibitor AMGMDS2. |
| | TP53 | Mutation – missense p.Glu286Lys exon 8/11 | Evidence A<br>Pathogenic<br>Likely pathogenic<br>Likely oncogenic | Resistance/Reduced Sensitivity:<br>- Rebemadlin.<br>- MDM2 inhibitor AMGMDS2. |
| | KRAS | Mutation – missense p.Thr20Ala exon 2/5 | Evidence A<br>Poor outcome | Sensitivity/Response:<br>- Trametinib.<br>- Binimetinib.<br>- Cobimetinib.<br>- AZD5438.<br>- GDC-0623. |
| | KRAS | Mutation – missense p.Leu19Phe exon 2/5 | Evidence A<br>Oncogenic<br>Poor outcome | Sensitivity/Response:<br>- Trametinib.<br>- Binimetinib.<br>- Cobimetinib.<br>- AZD5438.<br>- GDC-0623. |
| | NRAS | Mutation – missense p.Gln61Lys exon 3/7 | Evidence A<br>Oncogenic | Sensitivity/Response:<br>- Trametinib.<br>- Metformin. |
| | PIK3CA | Mutation – missense p.Ala1066Val exon 21/2 | Evidence A<br>Likely oncogenic | Sensitivity/Response:<br>- Alpelisib.<br>- Capivasertib.<br>- RLY-2608. |
| | PTEN | Mutation – frameshift | Evidence A<br>Pathogenic | Sensitivity/Response:<br>- Ipatasertib. |

| | | | | |
|---|---|---|---|---|
| | | p.Lys267ArgfsTer9 exon 7/9 | | - GSK26364771.<br>- AZD8186. |
| | NF1 | Mutation – frameshift p.Tyr930PhefsTer8 exon 21/5 | Evidence B | Sensitivity/Response:<br>- Cobimetinib.<br>- Trametinib. |

Note: It is important to underline an important number of alterations without a specific predictive value, but with clear functional relevance evidence of the genes APC, MSH6, MLH1, MUTYH, CDKN, CCND1, CCND3, FAS, FGF, RAC1, JAK2, HRAS, CTNNA1, RICTOR, RB1, and CIC.

**Supplementary Table 6** - Actionable gene variants (distributed per ESCAT evidence tier) found for patients included in C4.

For each variant, gene identification, nature of the alteration, functional relevance evidence for the alteration (A – Curated; B – Assumed; C - Predicted) and the predictive value of the alteration is provided (2 - Investigational; 3 – Hypothetical target: Alteration-drug match is associated with antitumor activity, but magnitude of benefit is unknown (potential cancer-repurposing opportunity); 4 – Hypothetical target: pre-clinical evidence of actionability).

| Tier | Gene | Alteration | Functional relevance evidence | Biomarker predictive value |
|---|---|---|---|---|
| **2** | MDM 2 | Copy number alteration (Amplification) | Evidence A<br>Oncogenic | Sensitivity/Response:<br>- Brigimadlin.<br>- Milademetan. |
| | TP53 | Mutation – missense p.Pro151Thr exon 5/11 | Evidence A<br>Pathogenic<br>Likely pathogenic<br>Likely oncogenic | Sensitivity/Response:<br>- Pazopanib.<br>- Vorinostat. |
| | TP53 | Mutation - stop gained p.Gln100Ter exon 4/11 | Evidence A<br>Pathogenic<br>Likely pathogenic | Sensitivity/Response:<br>- Pazopanib.<br>- Vorinostat. |
| | TP53 | Mutation – frameshift p.Arg213HisfsTer34 exon 6/11 | Evidence B | Sensitivity/Response:<br>- Pazopanib.<br>- Vorinostat. |
| **3** | MDM2 | Copy number alteration (Amplification) | Evidence A<br>Oncogenic | Sensitivity/Response:<br>- Brigimadlin.<br>- Ezabenlimab. |
| | TP53 | Mutation – missense p.Pro151Thr exon 5/11 | Evidence A<br>Pathogenic<br>Likely pathogenic<br>Likely oncogenic | Sensitivity/Response:<br>- Azacytidine.<br>- Eprenetapopt.<br>- Chemotherapy.<br>Resistance/Reduced Sensitivity:<br>- RG7112. |
| | TP53 | Mutation - stop gained p.Gln100Ter exon 4/11 | Evidence A<br>Pathogenic<br>Likely pathogenic | Sensitivity/Response:<br>- Azacytidine.<br>- Eprenetapopt.<br>- Chemotherapy.<br>Resistance/Reduced Sensitivity:<br>- RG7112. |
| | TP53 | Mutation – frameshift p.Arg213HisfsTer34 exon 6/11 | Evidence B | Sensitivity/Response:<br>- Azacytidine.<br>- Eprenetapopt.<br>- Chemoterapy. |

| | | | | |
|---|---|---|---|---|
| | | | | Resistance/Reduced Sensitivity:<br>- RG7112. |
| | MLH1 | Mutation – missense p.Arg385His exon 12/19 | Evidence C | Sensitivity/Response:<br>- Talazoparib. |
| | BARD1 | Mutation – missense p.Val713Met exon 11/11 | Evidence C | Sensitivity/Response:<br>- Olaparib. |
| **4** | CDK4 | Copy number alteration (Amplification) | Evidence A Oncogenic | Sensitivity/Response:<br>- Palbociclib.<br>- Abemaciclib. |
| | MDM2 | Copy number alteration (Amplification) | Evidence A Oncogenic | Sensitivity/Response:<br>- Brigimadlin.<br>- Milademetan. |
| | TP53 | Mutation – missense p.Pro151Thr exon 5/1 | Evidence A Pathogenic Likely pathogenic Likely oncogenic | Resistance/Reduced Sensitivity:<br>- Rebemadlin.<br>- MDM2 inhibitor AMGMDS2. |
| | TP53 | Mutation - stop gained p.Gln100Ter exon 4/11 | Evidence A Pathogenic Likely pathogenic | Resistance/Reduced Sensitivity:<br>- Rebemadlin.<br>- MDM2 inhibitor AMGMDS2 |
| | TP53 | Mutation – frameshift p.Arg213HisfsTer34 exon 6/11 | Evidence B | Resistance/Reduced Sensitivity:<br>- Rebemadlin.<br>- MDM2 inhibitor AMGMDS2. |

Note: It is important to underline an important number of alterations without a specific predictive value, but with clear functional relevance evidence of the genes SDHD, IGF1R, CALR, NTRK1.

**Supplementary Table 7 –** Characteristics of the studies that also employed unsupervised consensus clustering to analyze data originated from STS molecular profiling approaches.

This table provides, for each study, the STS histopathological subtypes of the samples that have been included, the types of molecular analyses that were performed (single or multi-omics, types of sequencing approaches that were used), the aims, methodological similarities and differences relative to our approach and, conceptually, the most relevant results.

| | **Samples** | **Molecular Analysis** | **Nature/ Aims** | **Methodological Approach** | **Results** |
|---|---|---|---|---|---|
| **Study Reference and Title** | | | | | |
| [20]<br><br>"Integrative Clustering Reveals a Novel Subtype of Soft Tissue Sarcoma With Poor Prognosis" | 247 STS samples<br><br>(including 56 DDLPS, 99 LMS,20 UPS) | Multi-omics: RNA and miRNA sequencing. | <u>Nature</u><br>Exploratory.<br><br><u>Aims</u><br>1. To evaluate a potential new STS classification.<br><br>2. To clarify putative molecular mechanisms behind each of the identified clusters. | <u>Similarities</u><br>Use of consensus clustering.<br><br><u>Dissimilarities</u><br>1. Combination of consensus clustering with similarity network fusion.<br><br>2. Construction of a competing endogenous RNA network based on differentially expressed mRNAs, lncRNAs and miRNA. | 1.Identification of 3 molecular clusters.<br><br>2. Correlation of one of the molecular clusters with worse prognosis.<br><br>3. Highlighting of a promising therapeutic target precisely for the cluster with the worse prognosis. |
| [41]<br><br>"Proteomic characterization identifies clinically relevant subgroups of soft tissue sarcoma" | 272 Chinese patients with an STS<br><br>(including 35 DDLPS, 52 LMS and 43 UPS patients) | Single-omics: Proteomics and phospho-proteomics<br><br>Both a proteomics (mass spectrometry-based) profiling and a phosphoproteomics (employing a Fe-NTA | <u>Aims</u><br>1. To unveil similarities and differences of the proteomic and phospho-proteomic profile of the included STS subtypes.<br><br>2. To uncover potential | <u>Similarities</u><br>Use of consensus clustering.<br><br><u>Dissimilarities</u><br>1. Concomitant use of 2 different types of unsupervised clustering (hierarchical and consensus) | 1. Similitude of proteomic characteristics between angiosarcoma and epithelial sarcoma (by hierarchical clustering).<br><br>2. Correlation of a high expression of SHC1 in angiosarcoma and |

| | | | | | |
|---|---|---|---|---|---|
| | | phosphopeptides enrichment technology) analysis were performed using samples (both tumor samples and matched tumor-adjacent tissues samples – to permit the study of the immune microenvironment). | mechanisms of STS metastasis.<br><br>3. To identify STS immune microenvironment features. | for different purposes. | epithelioid sarcoma with poor prognosis.<br><br>3. Identification of 3 proteomic clusters with various driven pathways and different clinical outcomes (by consensus clustering).<br><br>4. APEX1 and NPM1 promote, in the proteomic cluster portrayed by high cell proliferation rate, cell proliferation and drive the progression of cancer cells.<br><br>5. Highlighting of 3 immune subtypes with different tumor microenvironments (by consensus clustering).<br><br>6. Establishment of a potential association between immune evasion markers and metastasis development in STS. |

**Supplementary Table 8** – Most relevant distinctive molecular features of each transcriptomic cluster and conceptual rarity or novelty/originality of each feature.

| | **Distinctive Molecular Features** | **Rarity/ Originality** | **Additional Notes** |
|---|---|---|---|
| **Transcriptomic Cluster** | | | |
| **1** | 1. Over expression of CDK4 gene (possibly in correlation with an enrichment of this cluster in DDLPS, a subtype associated with CDK4 overexpression [34]).<br><br>2. Under expression of different genes (BRCA1, BRCA2, FANCD2, PALB2, RAD51, CHEK1, and BRIP1) involved in HRR. | <u>**Rarity**</u><br>1. Over expression of different genes involved in HRR in an STS cluster. Alterations in genes involved in the HRR genes is a relatively rare event in sarcomas. In a cohort of 7494 samples of soft tissue, bone and other sarcomas, only 2.5% of them harbored alterations in the homologous recombination repair pathways [34]. Another study reported that 4, 22, 21, 21 and 17% of sarcoma patients carry mutations in BRCA1, BRCA2, MDM2, PTEN and RAD1 respectively, and highlighted a subset of sarcomas that display high HRD [42].<br><br><u>**Originality**</u><br>1. Coexistence of CDK4 over expression and HRR genes under expression in an STS cluster. | The coexistence of these molecular features in this cluster may either represent an additive effect of the molecular contribution from sarcoma samples labelled as DDLPS (contributing with the CDK4 overexpression) and from sarcoma samples labelled as UPS and LMS (and also, potentially DDLPS) (contributing with the under expression of genes involved in HRR), or represent a new and previously undescribed sarcoma molecular subtype where these alterations concur. |
| **2** | <u>Over expression of cancer testis antigens (CTA).</u> | <u>**Rarity**</u><br>1. Over expression of CTA in a cluster composed by samples of specific histopathological | |

| | | |
|---|---|---|
| | 1. Over expression of MAGE (MAGE-A12, MAGE-A2B, MAGE-A3, MAGE-B1, MAGE-B2, MAGE-C2) genes.<br><br>2. Over expression of SSX (SSX-1, SSX-2, SSX-2B and SSX-3) genes. | subtypes (DDLPS, LMS, UPS). Typically, the STS histological subtypes in which a higher expression of CTA (specifically MAGE, SSX, NY-ESO and PRAME) is found are synovial sarcomas and myxoid round cell liposarcomas [43,44,45].<br><br>2. Over expression of SSX genes in an STS cluster composed by DDLMS, LMS and UPS samples.<br>The related SSX genes, SSX1 and SSX2, are fusion partners of SYT, integrating the synovial sarcoma pathognomonic translocation and the resulting fusion protein SYT-SSX (the expression (mRNA) of SYT-SSX is detected in 89-100% of either monophasic or biphasic synovial sarcoma cases) [45,46]. But SSX-genes may also be highly expressed in other STS subtypes, namely in LMS (mainly SSX-1), uterine LMS (SSX-1, SSX-2 and SSX-4), liposarcoma (mainly SSX-1, SSX-2 and SSX-3) and malignant fibrous histiocytoma (mainly SSX-2 and SSX-5) [46]. Interestingly, a significant fraction of STS samples co-express more than one SSX family member [46]. Another study reported a significant SSX mRNA expression in |

| | | |
|---|---|---|
| | | osteosarcoma, malignant peripheral nerve sheath tumor, malignant fibrous histiocytoma, liposarcoma, myxoid liposarcoma and LMS samples [47].<br><br>**<u>Originality</u>**<br>1. Over expression of MAGE-A12, MAGE-A2B, MAGE-A3, MAGE-B1, MAGE-B2, MAGE-C2 genes in an STS cluster. Among the genes that encode the MAGE-I family of CTA (consisting of MAGE-A, MAGE-B and MAGE-C), MAGE-A4 is the most commonly overexpressed gene in STS, being highly expressed in synovial sarcoma (90% mRNA and 82-83% protein levels) and myxoid/round cell liposarcoma (68% at the protein level) [44,45]. Other studies report a high expression of MAGE-A1,-A2 and -A3 in osteosarcomas, but not in any STS [44].<br><br>2. Co-over expression of MAGE genes (MAGE-A12, MAGE-A2B, MAGE-A3, MAGE-B1, MAGE-B2, MAGE-C2) different from the up mentioned MAGE-A4, and SSX-1, SSX-2, SSX-2B and SSX-3 genes. Co-expression of MAGE and SSX has already been reported in colorectal cancer (and was found to be directly |

| | | | |
|---|---|---|---|
| | | correlated with the development of metastasis to the liver [48]), but has never been clearly reported in the STS subtypes that compose our cohort. | |
| **3** | 1. Over expression of MHC class II/ HLA class II (HLA-DMA, HLA-DMB, HLA-DOA, HLA-DQA, HLA-DRA and HLA-DRB1) genes.<br><br>2. Under expression of CDKN (CDKN1C and CDKN2A) genes.<br><br>3. Under expression of FGFR (FGFR2 and FGFR3) genes. | **<u>Originality</u>**<br>1. Over expression of different genes that encode MHC/HLA class II peptides in STS.<br>In STS, the over expression of HLA class I genes (namely HLA-A) has been described in biphasic components of biphasic synovial sarcoma [49]. Another study looked into the genomic data of the neoplasms of 576 pediatric patients (262 of them with soft-tissue and bone sarcomas (rhabdomyosarcoma; non-rhabdomyosarcoma soft-tissue sarcoma; osteosarcoma; Ewing sarcoma)) with recurrent or refractory solid cancers enrolled in the MOSCATO-01 and the MAPPYACTS trials. This study unraveled a high HLA class I antigen expression in 27.1% of sarcoma samples, elevated frequencies of high HLA class I-positive samples in Ewing sarcoma, and osteosarcoma samples, high HLA-DR expression only in single specimens of osteosarcoma and an absence of HLA-DR expression in 73.4% of | The over expression of genes that encode different HLA class II peptides is a new finding in STS populations and may identify an STS cluster potentially more densely infiltrated by TIL, more prone to form TLS and characterized by enhanced antitumor immunity. |

| | | |
|---|---|---|
| | | all sarcoma samples, more prominently in rhabdomyosarcoma samples [50]. Indeed, the over expression of genes that encode HLA class II peptides in STS has not been previously reported. The expression of HLA Class II peptides has been described in an array of human neoplasms and despite being constitutively expressed on professional antigen-presenting cells (pAPCs) (such as dendritic cells, macrophages and B cells), they may also be expressed either by other cell types in the tumor microenvironment (such as antigen-presenting cancer-associated fibroblasts and lymphatic endothelial cells) or by tumor cells (the so-called tumor-specific HLA-II (tsHLA-II) [51]. Different studies in a variety of cancer types highlighted a correlation between high expression of tsHLA-II and favorable prognosis (improved PFS and OS), and between high expression of tsHLA-II and increased levels of both CD4+ and CD8+ tumor-infiltrating lymphocytes (TIL), absence of lymphovascular invasion, increased formation of tertiary lymphoid structures (TLS), upregulation of genes associated with |

| | | | |
|---|---|---|---|
| | | IFNg pathway activation, and higher levels of a plethora of different cytokines [51].<br><br>2. Under expression of FGFR genes in an STS cluster.<br>Aberrations in components of the FGFR signaling pathway have been highlighted in an array of different sarcoma subtypes, most notably gastrointestinal stromal tumors, rhabdomyosarcomas, and liposarcomas [52]. These alterations comprise genetic events such as translocations, mutations, and amplifications as well as transcriptional overexpression, but lead, in the great majority of the cases, to overexpression and not under expression of FGFR [52].<br><br>3. Coexistence of over expression of HLA class II genes, and under expression of CDKN2A and FGFR genes in an STS cluster. | |
| **4** | 1. Over expression of CLDN 4 gene.<br><br>2. Over expression of genes that encode different structural proteins. | **<u>Originality</u>**<br>Claudins are a multigene family of proteins that are crucial elements of epithelial cell tight junctions, usually mediating cell–cell adhesion and selectively | In ovarian cancer, the over expression of the claudin 4 is associated with worse prognosis (shorter OS), even though it does not impact the degree of |

| | | | |
|---|---|---|---|
| | | permitting the paracellular flux of ions and small molecules between cells [54]. They were recently brought to the spotlight as claudin 18.2, typically overexpressed in a subset of gastroesophageal neoplasms, became a compelling target for the either monoclonal antibodies (such as Zolbetuximab), whose use has provenly shown efficacy and survival advantage in phase III trials (such as SPOTLIGHT and GLOW), and also CAR-T cells and antibody-drug conjugates, whose use is currently being tested purely in an investigational context [54]. Claudin 4, more specifically, is typically over expressed in different epithelial malignancies types (namely breast, ovarian, lung, cervical, prostate, gastric and colorectal cancers), although its increased expression has not been previously described for any type of mesenchymal malignancy like STS (which is expected, considering that they are part of structures typically found in epithelia) [55]. | sensitivity to platinum [56].<br>In line with this findings, flourishing and prolific investigation on molecular therapies targeting claudin 4 has emerged, namely anti-claudin 4 extracellular domain antibodies, claudin 4 gene knockdown, clostridium perfringens enterotoxin (CPE), and C-terminus domain of CPE (C-CPE) [57].<br><br>This cluster is molecularly enriched in features that may reflect an increased transmembrane transportation and cell proliferative activity, which may characterize this molecular subset of STS. |
